# Prediction of Covid-19 vaccine effectiveness in adult populations and in clinically-vulnerable subgroups

**DOI:** 10.1101/2022.11.22.22282637

**Authors:** Oleg Volkov, Svetlana Borozdenkova, Alexander Gray

**Affiliations:** Xitific LTD, London, UK; IDEAPharma, Milton Keynes, UK

**Author notes:** Corresponding author: Oleg Volkov.

**Keywords:** Comirnaty (BNT162b2) and Vaxzevria (ChAdOx1 nCoV-19) Vaccines, Omicron and Earlier Variants SARS-CoV-2, Prediction of Covid Vaccine Effectiveness, Neutralising Antibody Titres, Legacy Immunogenicity Study

## Abstract

Predictions of Covid vaccine effectiveness could support rapid and effective measures against the pandemic. Our modelling boosts the accuracy and applications of these predictions, especially to subgroups. We model the symptomatic effectiveness of Comirnaty or Vaxzevria with 50% neutralising antibody titres from a large UK immunogenicity study and with up to 68 effectiveness estimates from 23 vaccine studies. We predicted effectiveness in adult populations, age and disease subgroups, with 45% (95% CI: 27–63) predicted against Omicron BA.1 for Comirnaty boosters in haemodialysis patients. Prediction errors for two Comirnaty doses in adults were 1.9%, 2.6% and 0.4%, against the Alpha, Beta and Delta variants, versus 3.6%, 28% and 8.7% with a state-of-the-art alternative; and for Vaxzevria, 1.1% and 0.7% against Alpha and Delta, versus 18% and 20.4%. Identical titres implied between 18% (95% CI: 1–33) and 31% (95% CI: 13–50) lower Comirnaty effectiveness against Omicron BA.1 than Delta.

---

After three years living with Covid-19, the public has little appetite for pandemic measures. Yet, SARS-CoV-2 keeps evolving and causing or contributing to deaths — including 22,143 last August in the US alone[1]. Especially now, pandemic measures must be effective, non-disruptive and least costly. And for this, they must reflect vaccine effectiveness: underestimated VE can lead to rushed vaccine boosters and thus to wasted resources, vaccine toxicities and hesitancy; overestimated VE, to delayed boosters, prematurely abandoned masks and thus to preventable hospitalisations and deaths. Estimating VE in observational studies, however, takes weeks or months, and before estimates are ready, the next variant/subvariant may become prevalent. In contrast, VE could be predicted cheaply and rapidly from correlates of vaccine protection (CoPs), such as levels of vaccine-induced antibodies or another biomarker. *Definitive* CoPs have been established for Vaxzevria[2], mRNA-1273[3] and Ad26.COV2.S[4] Covid vaccines in randomised controlled trials, and for Vaxzevria and Comirnaty[5, 6], in observational VE studies. As a human study is costly and time-consuming, these CoPs have limited scope and precision: they were investigated for one dose[4] or two doses[2, 3, 5, 6] only, and many of them are highly uncertain outside mid-range antibody levels (Figure 4 of 2, 3, 6). Established for one variant[2, 3, 5, 6] or several jointly[4], CoPs may not apply to a new variant, and more studies could be needed[6]. Alternatively, *data-driven* CoPs, which exploit statistical associations between VE and biomarker levels, can be quickly developed without large additional studies. For example, [7] used genetic distances between a variant and the ancestral SARS-CoV-2 to predict VE in twice-vaccinated adults. As such distances lack a clear, easy-to-test dependence on vaccination parameters — doses, subgroup factors, time post-dose and the like — VE prediction for boosters, subgroups, etc. would be challenging.

We consider predicting VE from data-driven CoPs based on vaccine-induced antibody levels — an approach pioneered[8, 9] for vaccine efficacy against the ancestral Wild Type and extended[10, 11] to variants; see Discussion for details. Although these predictions could be very valuable — for instance, when definitive CoPs or reliable VE estimates are unavailable — issues with accuracy, validation and support for key applications limit their policy use. By addressing these issues we aim to widen the applications and impact of VE predictions. Our main target is VE in subgroups, such as cancer patients aged over 50, defined by demographic, clinical or combined factors. Learning VE is crucial in protecting the immunocompromised: a fraction of the population, they comprise over 40% of hospitalisations with breakthrough Covid[12] but currently are “largely forgotten and left to their own devices to manage risk.”[13] As young healthy adults have strong immunity, but also high rates of vaccine toxicity and hesitancy, learning VE for them could support less frequent boosting. Observational VE studies, however, are slow or unfeasible in a smaller subgroup, especially one with extra shielding and few unvaccinated members. Data-driven prediction is also limited[14] — we only found ref[11] with two age subgroups. We also target prediction for partial vaccination and seek more accurate VE predictions overall.

We developed a data-driven VE framework, by building upon[9, 10], and apply it here to the symptomatic VE of Comirnaty or Vaxzevria. It models VE as a function of vaccine-induced 50% neutralising antibody titres (NAbTs) against a variant/subvariant, which correlate with the effectiveness and efficacy of Covid vaccines[2, 3, 4, 9, 8, 10]. Each model combines NAbTs and VEs for several variants, doses and post-dose intervals, for one or more vaccines. We require comparable NAbTs, such as obtained in a single large study or in several similar studies. Here we use NAbTs from the Legacy longitudinal immunogenicity study, run in the UK by the University College London Hospitals and the Francis Crick Institute[15, 16, 17]. The “Primary” Legacy[15, 16] enrolled once or twice-vaccinated adults — 346 with Comirnaty and 113 with Vaxzevria — and investigated an ancestral strain with the D614G mutation, and the Alpha, Beta and Delta variants. The “Post-Primary” Legacy[17] had similar participants after two doses or boosters and assessed the Omicron BA.1 subvariant versus Alpha and Delta. The disease subgroup examples use NAbTs from Legacy-affiliated studies[18, 19] in patients on haemodialysis.

“Primary” VE models were fitted for the pre-Omicron variants by using 43 symptomatic VE estimates, in adults, from fifteen Comirnaty studies and/or 25 from eight Vaxzevria studies. Predictions against a pre-Omicron variant were compared with state-of-the-art predictions[10]. The “Primary” Comirnaty model was applied to predict VE against Delta or Omicron BA.1 from “Post-Primary” Legacy NAbTs[17]; we also fitted an individual VE model for either variant. Our results on BA.1 could apply to later subvariants: symptomatic VE against BA.2 is similar[20, 21], while VE against hospitalisation with BA.4 and BA.5 is similar to that with BA.2[22].

## Results

We first introduce and validate our framework for primary vaccination — either partial or full — and the four pre-Omicron variants in “Primary” Legacy[15, 16].

### Fitted models

We implement direct VE modelling for each vaccine individually; indirect modelling of Vaxzevria VE with a model fitted with Comirnaty data and evaluated at Vaxzevria NAbTs; and combined modelling for both vaccines. (See Methods for modelling details and Supplemental results for parameter estimates, sample sizes and data options in the fitted models.) The direct Comirnaty model is plotted in Figure 1a versus log_10_ geometric mean titre (GMT) of a NAbT distribution per dose and variant combination. This model is used for indirect modelling of Vaxzevria VE, and is shown in Figure 1b together with the direct Vaxzevria model. The fitted combined model is shown in Figure 1c. Each model displays a strong relationship between NAbTs and VE estimates, which is roughly linear on a log scale. Vaxzevria modelling is based on fewer VE estimates (25 v 43) and Legacy unique participants (106 v 175) than Comirnaty modelling, while the median age, of 36, in the Vaxzevria cohort is much below that of Vaxzevria vaccinees in general. Yet, both the direct and indirect Vaxzevria models fit well: the latter deviates from the former at the dose/variant combinations used by between −4.1% and 4.2%, where “−” means under-prediction. This result and the good combined fit in Figure 1c suggest a common trend for both vaccines, despite their distinct underlying cohorts and platforms, mRNA versus adenovirus vector. The 95% prediction bands cover approximately 93% estimates for the Comirnaty and combined model fits, and 96% for the direct Vaxzevria model fit. (Five out of eight efficacy estimates in Figure 1a of [9] are inside the 95% prediction bands, giving a coverage of 63%.)

**Figure 1:**
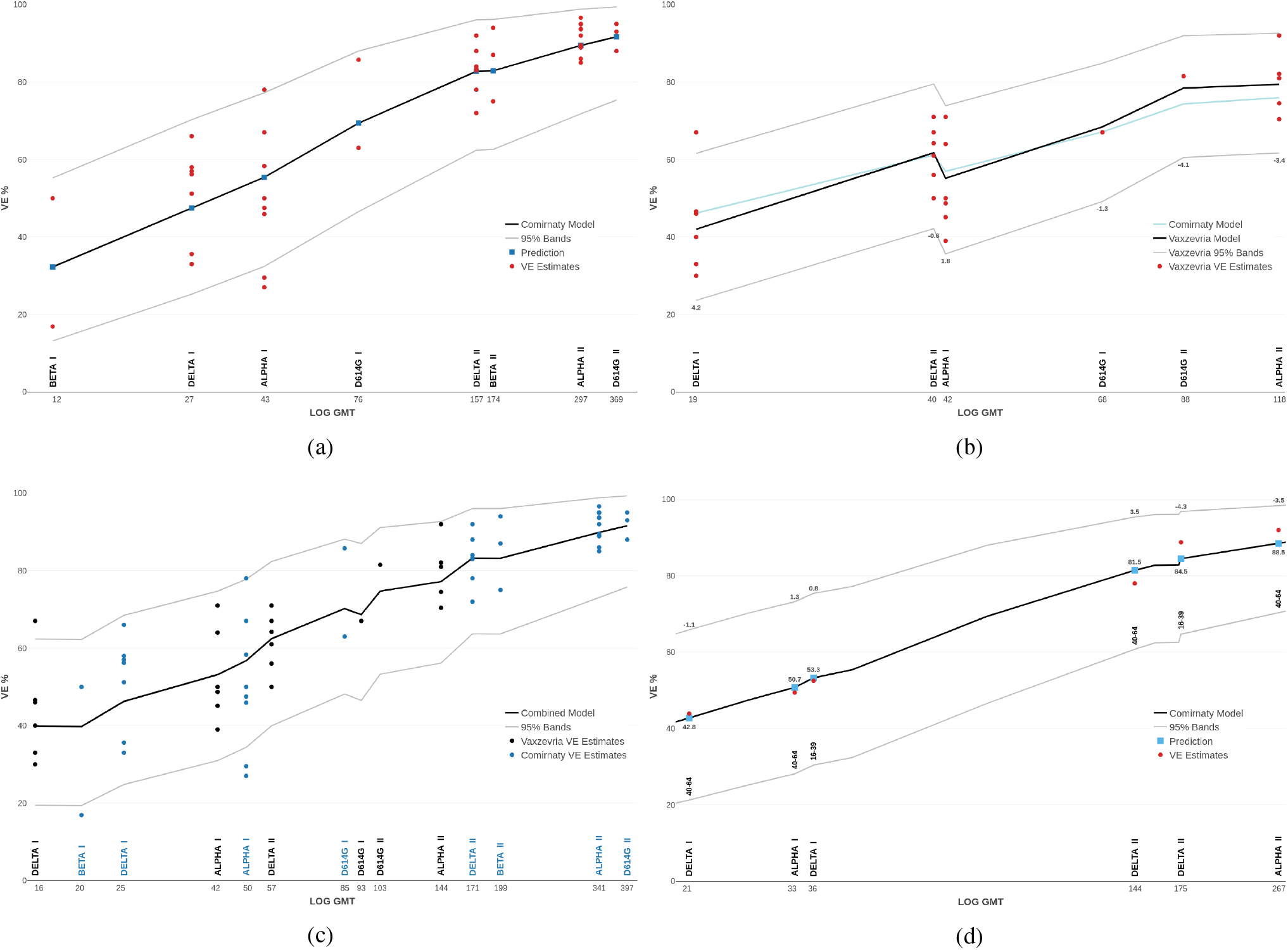
Fitted VE models (2) in adult populations versus log_10_ GMT, which is labelled on the original scale. As GMT is a scalar summary of NAbTs, each plot is a simplification; see Section A.2. Predictions and VE estimates are per variant and dose (a Roman numeral). (a) Comirnaty VE: direct model. (b) Vaxzevria VE: direct model and indirect model from Comirnaty data, and their prediction differences. (c) Combined VE model fitted to data on both vaccines. (d) Comirnaty model (a) predicts Comirnaty VE against Alpha or Delta in the age subgroups indicated; the GMT is shown for the age subgroups of Legacy; VE estimates are from ref[23]; prediction deviations from them are shown above the prediction band.

### Age subgroups

Predicting VE is straightforward for demographic subgroups that are well-represented by Legacy participants. Here, we predict Comirnaty VE against Alpha or Delta for a younger 16–39 and older 40–64 age subgroups, for whom VE is reported in [23]; see Table 4. We first select seronaive participants of Legacy[15] in these age ranges: the younger subgroup with 54 and 46 Comirnaty vaccinees after one dose and two doses, respectively; and the older subgroup with 72 and 61. To predict subgroup VE, we input their NAbTs into the previous Comirnaty model, in Figure 1a. The predictions, in Figure 1d, deviate by between −4.3% and 3.5% from the VE estimates[23] and are quite accurate, despite the demographic differences between the VE study population and the Legacy cohorts, with no-one younger than 20.

### Diabetes and kidney disease subgroups

Our framework applies to subgroups whose NAbTs are studied outside of Legacy but under similar experimental protocols. To predict VE we input subgroup NAbTs into a model fitted to Legacy data; here it is the Comirnaty model in Figure 1a. These NAbTs must be compatible with Legacy NAbTs — in particular, between-study error should be known or negligible, as we assume. The current example uses the study[18] in long-term haemodialysis patients, whose post-vaccination NAbTs were determined identically to Legacy NAbTs. We focus on the study’s 55 seronaive participants after two Comirnaty doses, 38% of whom were female, and whose mean age (standard deviation) was 64 (12). Since no symptomatic VE estimates could be found for haemodialysis populations, we predict VE separately for 25 patients with diabetes and 30 with non-diabetic kidney disease. These predictions are compared to published VE estimates in diabetes and kidney disease populations; see Figure 2. As these estimates are not variant-specific, we predict combined VE for the variants prevalent in a given VE study; see Methods. The VE predictions and estimates in the disease subgroups are lower than the corresponding VE estimates in healthy adults. The prediction and estimate per study are reasonably close, with the largest absolute error being 7% and the smallest, 2%. This is despite the small subgroups studied[18] and potentially large differences — including in disease characteristics — between them and the disease subgroups in the VE studies. As haemodialysis usually starts at late-stage kidney dysfunction, the participants[18] are probably more impaired and older than diabetes or kidney disease patients in general (although a shorter life expectancy under haemodialysis might offset the age difference).

**Figure 2:**
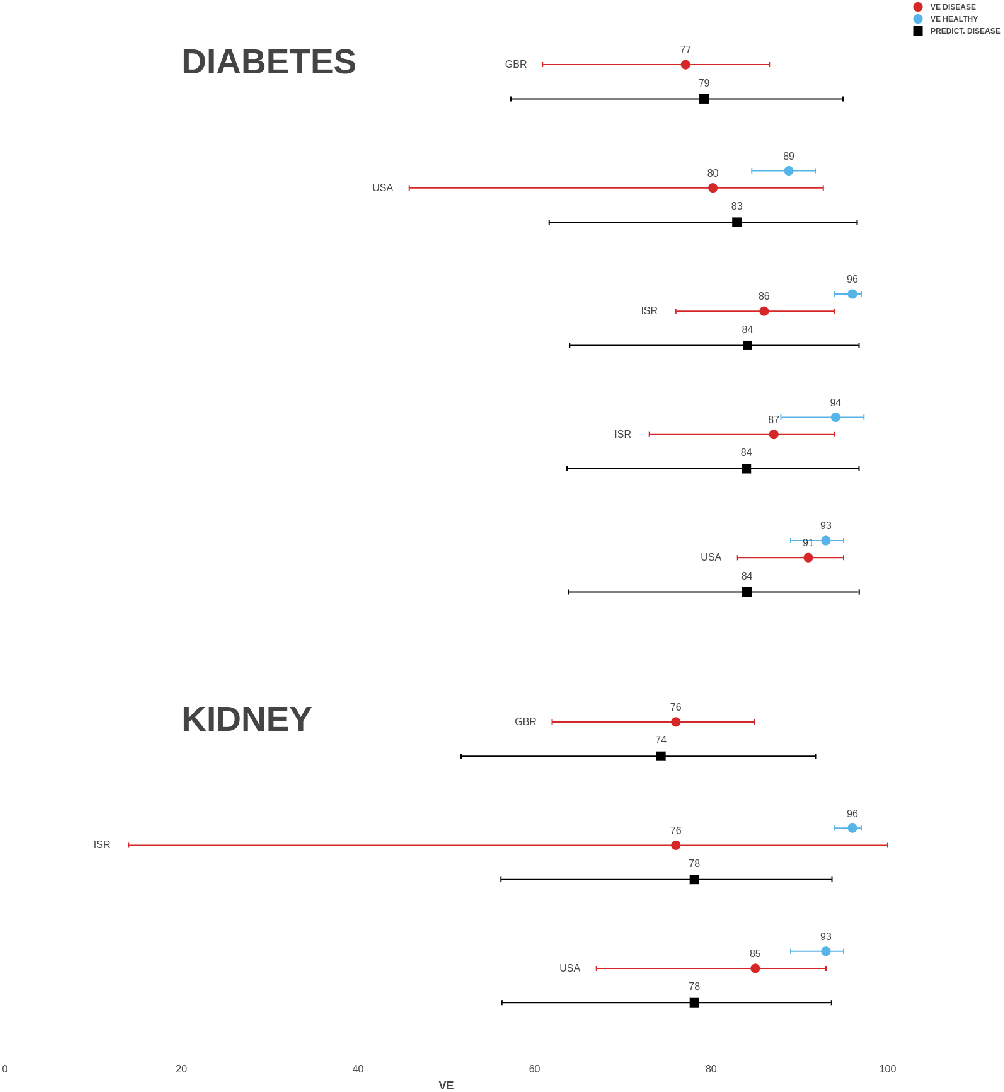
Two-dose Comirnaty VE, in %, predicted in diabetes and chronic kidney disease subgroups. Also shown are VE estimates in subgroups and in the healthy adult population of the same study.

### Predictions per variant

We obtain “leave-one-variant-out” predictions of VE against a variant by fitting a model without this variant. These predictions are compared to the average of at least two VE estimates reported per vaccine, dose and variant; see Tables 2 and 3. For Vaxzevria, both direct and indirect (Comirnaty-based) models are fitted without the variant and evaluated at Vaxzevria NAbTs for the variant. For one dose, the deviations, in %, from average VE estimates are 2.9_*α*_ and −5.2_*δ*_ for direct prediction and 5.8_*α*_ and −2.4_*δ*_ for indirect prediction, where the subscript denotes the variant; see Figure 9. For two doses, these deviations are −1.1_*α*_ and −0.7_*δ*_ for direct prediction, and −3.3_*α*_ and −3.5_*δ*_ for indirect prediction; see Figure 3a. The 95% prediction bands cover VE estimates well, especially for two doses.

**Table 1:** VE is predicted from the separately-fitted Delta and Omicron models evaluated at Omicron NAbTs for adult groups[17] and haemodialysis (HD) patients[19]. The predicted VE differences between the Delta and Omicron models are based on 100,000 simulations from pairs of predictive Beta distributions per group. Each confidence interval (CI) for GMT is based on 100,000 bootstrap replications of reported Omicron NAbTs[17, 19].

| Group | Omicron NAbTs |  | Delta Model |  | Omicron Model |  | VE Difference |  |
| --- | --- | --- | --- | --- | --- | --- | --- | --- |
|  | GMT | 95% CI | VE % | 95% CI | VE % | 95% CI | % | 95% CI |
| Early II | 82 | [67–98] | 65 | [59–71] | 36 | [19–54] | 29.0 | [9.7–46.4] |
| Late II | 24 | [18–31] | 39 | [33–45] | 21 | [8–38] | 18.2 | [0.6–32.5] |
| Boost III | 304 | [240–364] | 87 | [83–91] | 56 | [38–74] | 30.8 | [12.8–49.6] |
| HD Boost III | 119 | [52–251] | 66 | [60–72] | 45 | [27–63] | 21.4 | [2.1–40.0] |

**Table 2:**
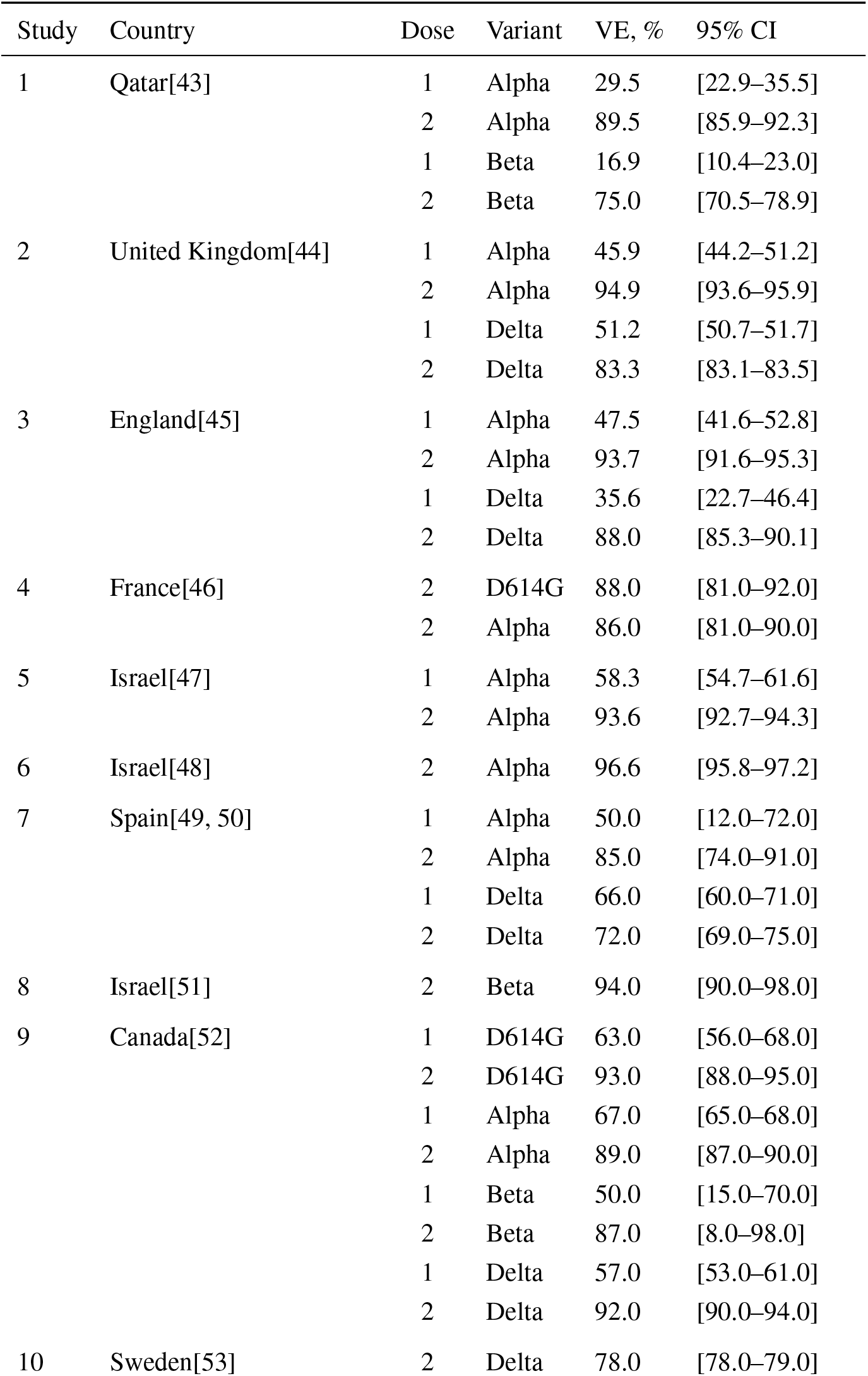
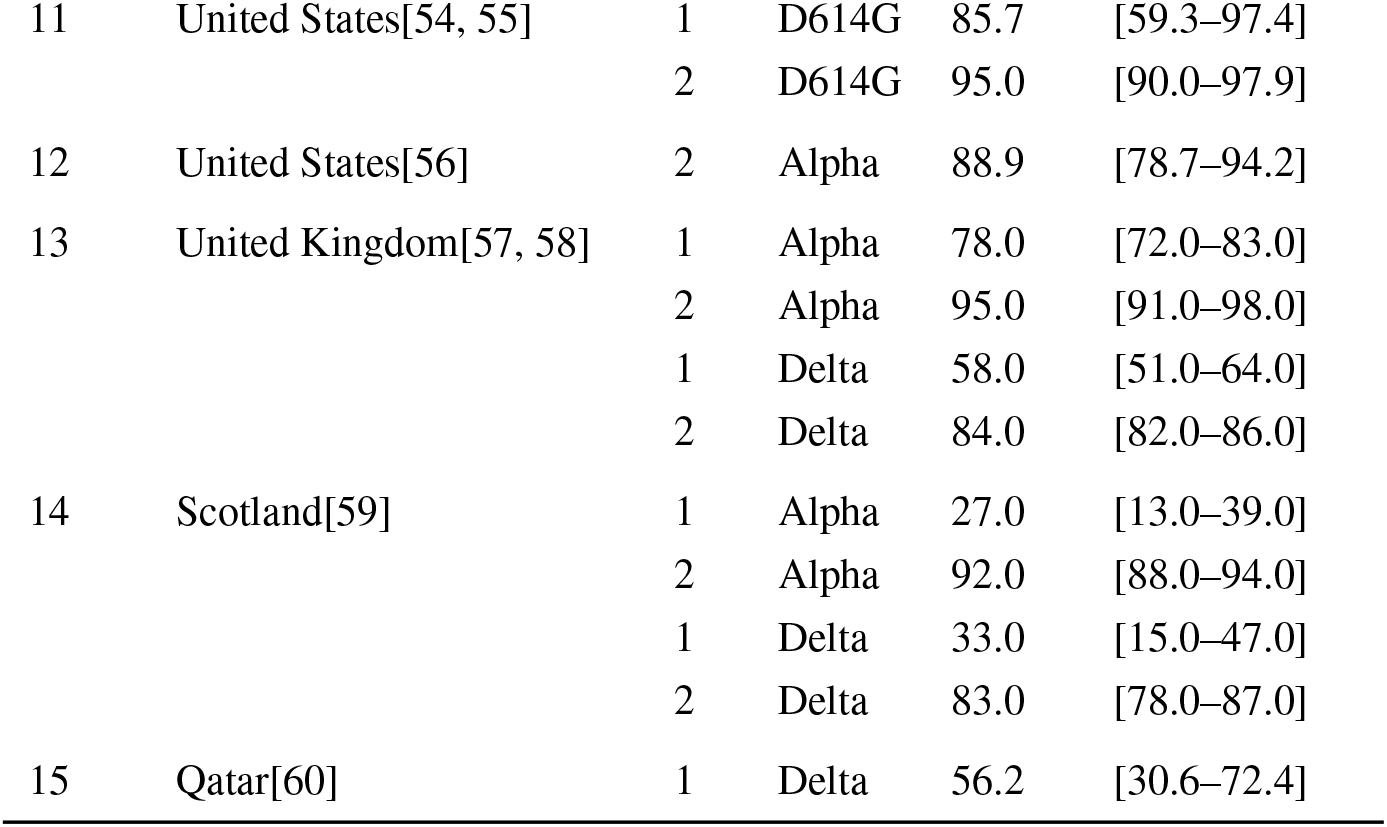
Studies of symptomatic VE of Comirnaty in adult populations.

| Study | Country | Dose | Variant | VE, % | 95% CI |
| --- | --- | --- | --- | --- | --- |
| 1 | Qatar[43] | 1 | Alpha | 29.5 | [22.9–35.5] |
|  |  | 2 | Alpha | 89.5 | [85.9–92.3] |
|  |  | 1 | Beta | 16.9 | [10.4–23.0] |
|  |  | 2 | Beta | 75.0 | [70.5–78.9] |
| 2 | United Kingdom[44] | 1 | Alpha | 45.9 | [44.2–51.2] |
|  |  | 2 | Alpha | 94.9 | [93.6–95.9] |
|  |  | 1 | Delta | 51.2 | [50.7–51.7] |
|  |  | 2 | Delta | 83.3 | [83.1–83.5] |
| 3 | England[45] | 1 | Alpha | 47.5 | [41.6–52.8] |
|  |  | 2 | Alpha | 93.7 | [91.6–95.3] |
|  |  | 1 | Delta | 35.6 | [22.7–46.4] |
|  |  | 2 | Delta | 88.0 | [85.3–90.1] |
| 4 | France[46] | 2 | D614G | 88.0 | [81.0–92.0] |
|  |  | 2 | Alpha | 86.0 | [81.0–90.0] |
| 5 | Israel[47] | 1 | Alpha | 58.3 | [54.7–61.6] |
|  |  | 2 | Alpha | 93.6 | [92.7–94.3] |
| 6 | Israel[48] | 2 | Alpha | 96.6 | [95.8–97.2] |
| 7 | Spain[49, 50] | 1 | Alpha | 50.0 | [12.0–72.0] |
|  |  | 2 | Alpha | 85.0 | [74.0–91.0] |
|  |  | 1 | Delta | 66.0 | [60.0–71.0] |
|  |  | 2 | Delta | 72.0 | [69.0–75.0] |
| 8 | Israel[51] | 2 | Beta | 94.0 | [90.0–98.0] |
| 9 | Canada[52] | 1 | D614G | 63.0 | [56.0–68.0] |
|  |  | 2 | D614G | 93.0 | [88.0–95.0] |
|  |  | 1 | Alpha | 67.0 | [65.0–68.0] |
|  |  | 2 | Alpha | 89.0 | [87.0–90.0] |
|  |  | 1 | Beta | 50.0 | [15.0–70.0] |
|  |  | 2 | Beta | 87.0 | [8.0–98.0] |
|  |  | 1 | Delta | 57.0 | [53.0–61.0] |
|  |  | 2 | Delta | 92.0 | [90.0–94.0] |
| 10 | Sweden[53] | 2 | Delta | 78.0 | [78.0–79.0] |
| 11 | United States[54, 55] | 1 | D614G | 85.7 | [59.3–97.4] |
|  |  | 2 | D614G | 95.0 | [90.0–97.9] |
| 12 | United States[56] | 2 | Alpha | 88.9 | [78.7–94.2] |
| 13 | United Kingdom[57, 58] | 1 | Alpha | 78.0 | [72.0–83.0] |
|  |  | 2 | Alpha | 95.0 | [91.0–98.0] |
|  |  | 1 | Delta | 58.0 | [51.0–64.0] |
|  |  | 2 | Delta | 84.0 | [82.0–86.0] |
| 14 | Scotland[59] | 1 | Alpha | 27.0 | [13.0–39.0] |
|  |  | 2 | Alpha | 92.0 | [88.0–94.0] |
|  |  | 1 | Delta | 33.0 | [15.0–47.0] |
|  |  | 2 | Delta | 83.0 | [78.0–87.0] |
| 15 | Qatar[60] | 1 | Delta | 56.2 | [30.6–72.4] |

**Table 3:** Studies of symptomatic VE of Vaxzevria in adult populations.

| Study | Country | Dose | Variant | VE, % | 95% CI |
| --- | --- | --- | --- | --- | --- |
| 1 | United Kingdom[44] | 1 | Alpha | 45.1 | [43.4–46.7] |
|  |  | 2 | Alpha | 82.1 | [79.4–84.5] |
|  |  | 1 | Delta | 46.6 | [45.8–47.5] |
|  |  | 2 | Delta | 64.2 | [63.9–64.5] |
| 2 | England[45] | 1 | Alpha | 48.7 | [45.2–51.9] |
|  |  | 2 | Alpha | 74.5 | [68.4–79.4] |
|  |  | 1 | Delta | 30.0 | [24.3–35.3] |
|  |  | 2 | Delta | 67.0 | [61.2–71.8] |
| 3 | United Kingdom[65] | 2 | D614G | 81.5 | [67.9–89.4] |
|  |  | 2 | Alpha | 70.4 | [43.6–84.5] |
| 4 | Spain[49, 50] | 1 | Alpha | 50.0 | [37.0–61.0] |
|  |  | 1 | Delta | 46.0 | [37.0–54.0] |
|  |  | 2 | Delta | 56.0 | [48.0–63.0] |
| 5 | Canada[66] | 1 | D614G | 67.0 | [38.0–82.0] |
|  |  | 1 | Alpha | 64.0 | [60.0–68.0] |
|  |  | 1 | Delta | 67.0 | [44.0–80.0] |
| 6 | Sweden[53] | 2 | Delta | 50.0 | [41.0–58.0] |
| 7 | United Kingdom[57, 58] | 1 | Alpha | 71.0 | [62.0–78.0] |
|  |  | 2 | Alpha | 92.0 | [78.0–97.0] |
|  |  | 1 | Delta | 40.0 | [28.0–50.0] |
|  |  | 2 | Delta | 71.0 | [66.0–74.0] |
| 8 | Scotland[59] | 1 | Alpha | 39.0 | [32.0–45.0] |
|  |  | 2 | Alpha | 81.0 | [72.0–87.0] |
|  |  | 1 | Delta | 33.0 | [23.0–41.0] |
|  |  | 2 | Delta | 61.0 | [51.0–70.0] |

**Figure 3:**
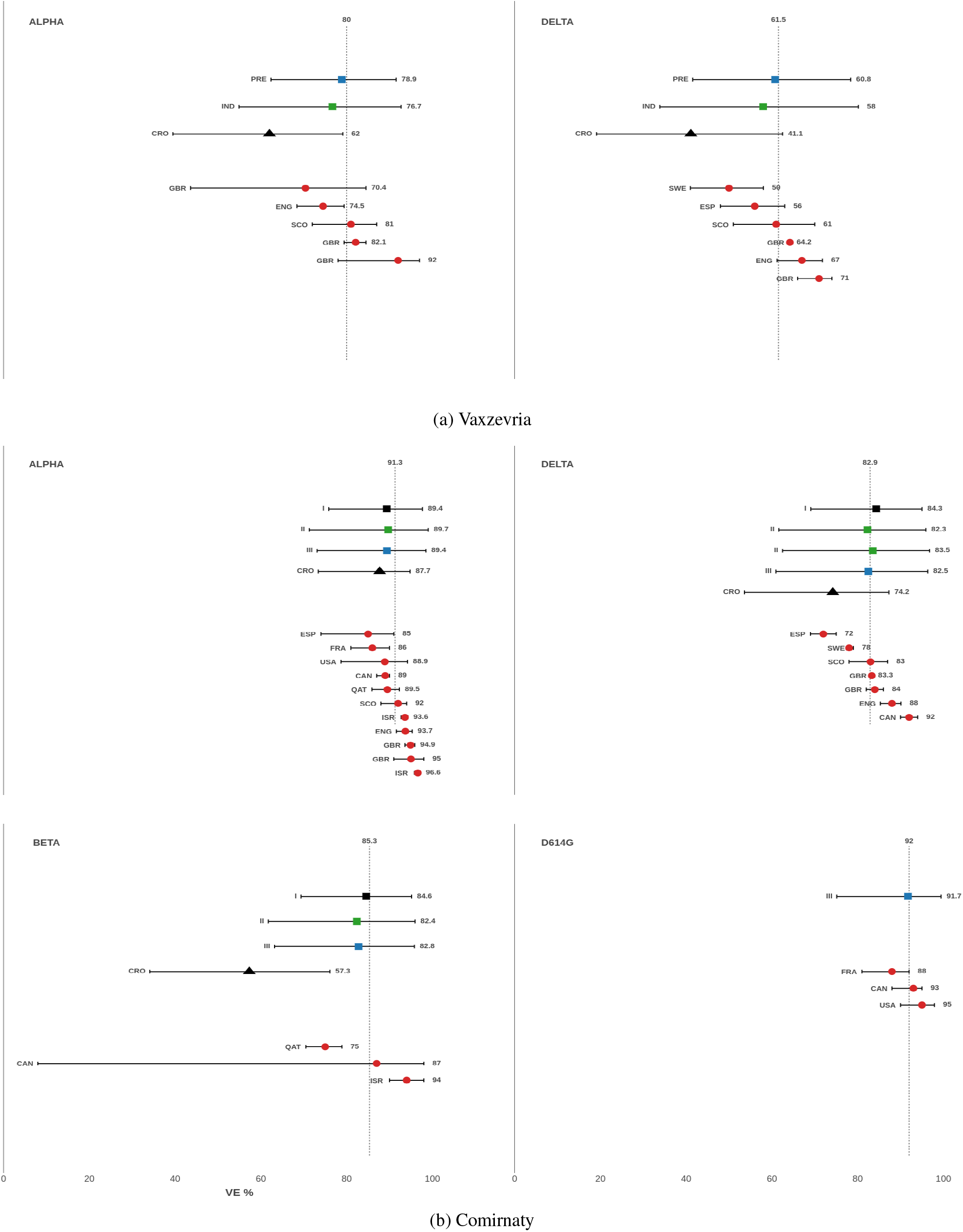
Predictions of two-dose VE against a variant versus VE estimates from indicated countries; see Tables 2 and 3. (a) “Leave-one-variant-out” predictions of Vaxzevria VE, including PRE direct predictions from Vaxzevria data; IND indirect prediction from a Comirnaty-fitted model at Vaxzevria NAbTs; and CRO predictions in [10].(b) Predicted Comirnaty VE. Epoch I: predictions from D614G data. Epoch II: predictions from D614G and another variant; Epoch III: predictions from three other variants, ie, “leave-one-variant-out” predictions; CRO predictions in [10].

**Figure 4:**
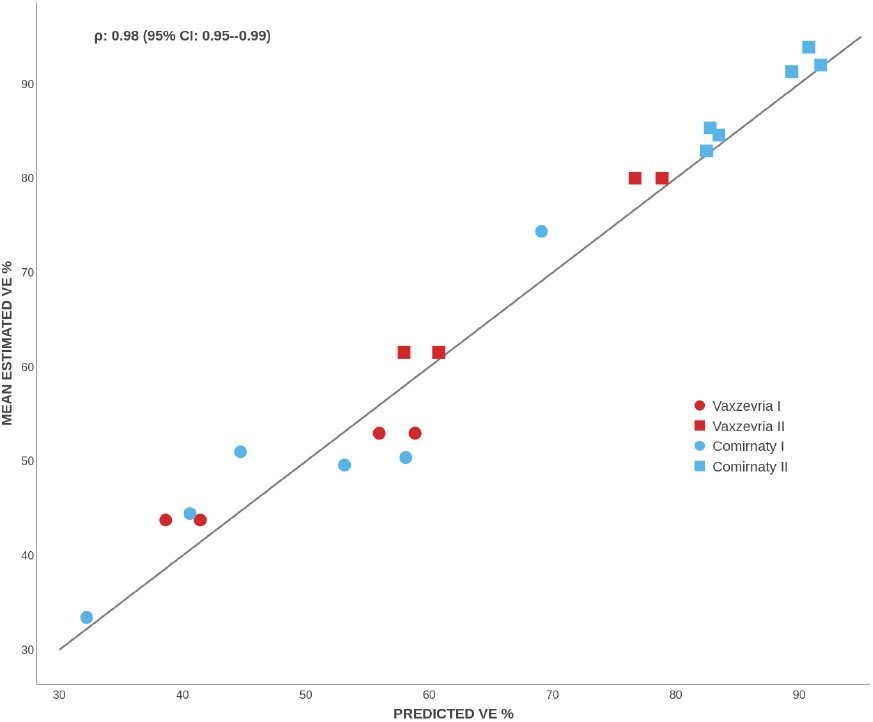
Leave-one-variant-out predictions from the direct and indirect Vaxzevria models and from direct Comirnaty model (including UK-only) versus mean VE estimates per vaccine, dose and variant. The estimated concordance coefficient *ρ* is displayed on top.

To assess prediction of VE as variants emerge or from only a few variants, we predict Comirnaty VE against successive variants from data on previous and concurrent variants. We define the following “epochs” after D614G dominance: *Epoch 1:* Either Alpha or Beta dominates a large territory, for instance, Alpha in Europe and Beta in South Africa. To predict VE against either variant, and also against Delta, we use data on D614G only. *Epoch 2:* VE estimates for Alpha and Beta become available, perhaps at different times. Therefore, we predict VE against Alpha from D614G and Beta; VE against Beta from D614G and Alpha; and VE against Delta from either of these pairs. *Epoch 3:* Delta becomes predominant, and VE against it can be predicted from the three previous variants. Here, we make such “leave-one-variant-out” predictions against each of the four variants. As illustrated in Figure 3b predictions for two doses are accurate already at Epoch 1. Although less accurate, predictions for one dose improve at later epochs with more data. The “leave-one-variant-out” predictions deviate from the average VE estimates by −5.3_*dg*_, 7.7_*α*_, −1.3_*β*_, −6.3_*δ*_ per cent after one dose (Figure 9) and by −0.3_*dg*_, −1.9_*α*_, −2.6_*β*_, −0.4_*δ*_ after two doses (Figure 3b). All the two-dose VE estimates are inside the corresponding 95% prediction bands.

We next predict Comirnaty VE for the UK adult population from UK-only data; such predictions could be useful for country-specific policies. Since after Comirnaty authorisation and before Omicron, only Alpha and Delta were prevalent in the UK, we use 16 estimates corresponding to these variants; see Table 2. “Leave-one-variant-out” predictions against either variant only use eight estimates against the other variant; see Figure 10. Despite the limited data, these predictions deviate from the UK-average VE estimates only by 3.5_*α*_, −3.9_*δ*_, and by −3.1_*α*_, −1.1_*δ*_ per cent for one dose and two doses, respectively.

For either vaccine, the predictions are close to the corresponding average VE estimates. The concordance coefficient[24, 25] for all the leave-one-variant-out and UK-only predictions combined is estimated as 0.98 (95% CI: 0.95–0.99); see Figure 4. The predictions are less accurate for one dose, perhaps due to more censored NAbTs and more uncertain VE estimates. Indeed, per-variant spreads between the smallest and largest estimates are greater after one dose by 1.7–4.4 times for Comirnaty and by 1.5–1.8 for Vaxzevria; see Figure 1c. Note that prediction variance in beta regression is maximal around 50% — and this is where the one-dose VE estimates cluster — which supports the beta distribution assumption for VE.

We now consider comparable VE predictions from the state-of-the-art paper[10]. These predictions, only available for two doses, were extracted from its Figure 2a with PlotDigitizer. (We omit predictions in [11], made for different post-dose periods.) The predictions against Alpha are accurate for Comirnaty VE but deviate from the average VE estimate by −18% for Vaxzevria; see Figure 3. Predicted Comirnaty VE against Beta deviates by −28%, versus −2.6% for the proposed prediction. Predicted Comirnaty VE against Delta deviates by −8.7% for Comirnaty, versus 0.4% for the proposed prediction. Predicted Vaxzevria VE against Delta deviates by −20.4%, versus −0.4% and −3.5% for the proposed direct and indirect predictions, respectively. The coverage of VE estimates by 95% prediction bands[10] is less accurate.

### Comirnaty VE against Delta or Omicron

We next consider the VE of full primary or of boosted vaccination with Comirnaty against the Delta variant or Omicron BA.1 subvariant. “Post-Primary” Legacy[17] tested Alpha, Delta and Omicron BA.1 in the following Comirnaty groups: “Early”, 2–6 weeks after two doses; “Late”, 12–16 weeks after two doses; and “Booster”, 2–6 weeks after three doses. Comirnaty VE is predicted here against Delta or Omicron BA.1 for these groups and post-dose periods, and compared with average VE estimates, based on Table 6. Suppose first that these estimates were unavailable for model fitting; hence, we predict VE by evaluating a “Primary” Comirnaty model at “Post-Primary” NAbTs. As assays and participants differed between the studies, their NAbTs had to be made comparable. As detailed in Methods, we divide all the Post-Primary NAbTs against Delta by 1.51 and those against Omicron by 1.6. The predictions against Delta are accurate, with % deviations of −4.2_*E*_, −5_*L*_ and −2.2_*B*_ for the indicated groups; Figure 5a. Some under-prediction is reasonable, as the Primary model assumed VE estimates over 3–5 months post-dose, unlike the shorter periods in Post-Primary Legacy. We emphasise over-predicted VE against Omicron, with % deviations of 17.5_*E*_, 26.2_*L*_ and 27.9_*B*_. Obtained for different post-dose periods, symptomatic VE predictions[26] over-predict Omicron VE by a similar magnitude.

**Table 4:** Comirnaty VE in age subgroups in a UK study[23]; Table S6. Prior Covid cases were excluded.

| Age | Dose | Variant | VE, % | 95% CI |
| --- | --- | --- | --- | --- |
| 40–64 | 1 | Alpha | 49.4 | [45.6–53.0] |
| 40–64 | 2 | Alpha | 92.0 | [86.7–95.2] |
| 16–39 | 1 | Delta | 52.5 | [52.1–53.0] |
| 16–39 | 2 | Delta | 88.8 | [88.6–89.0] |
| 40–64 | 1 | Delta | 43.9 | [41.9–45.8] |
| 40–64 | 2 | Delta | 78.0 | [77.5–78.5] |

**Table 5:**
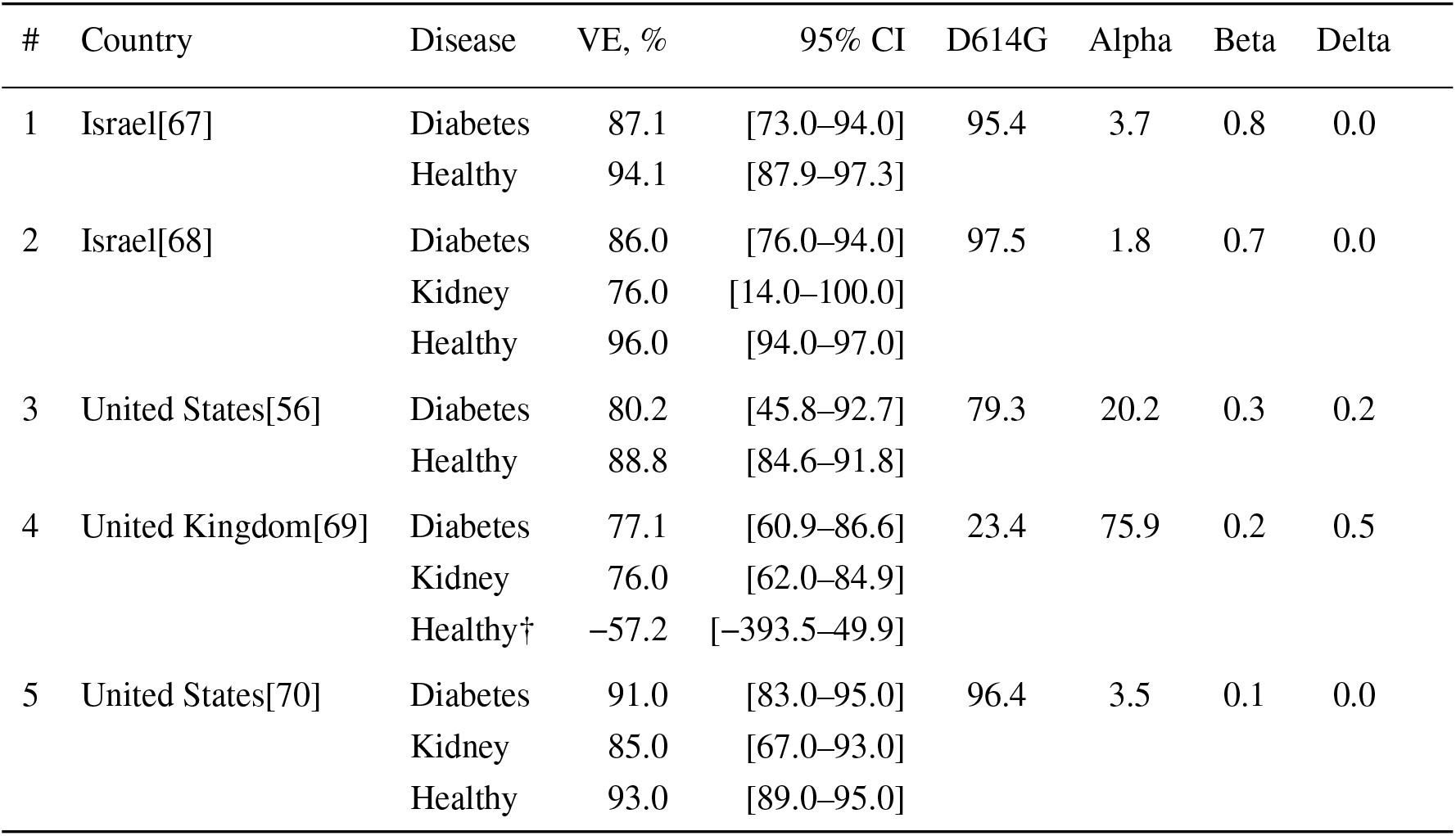
Studies of symptomatic VE of two Comirnaty doses in kidney/diabetes disease subgroups and in healthy subgroups of adult populations. Prior Covid cases are excluded. Approximated shares, in %, of Covid cases for the study country and dates are shown per variant. These estimates, except†, were used for comparison in Figure 2.

**Table 6:**
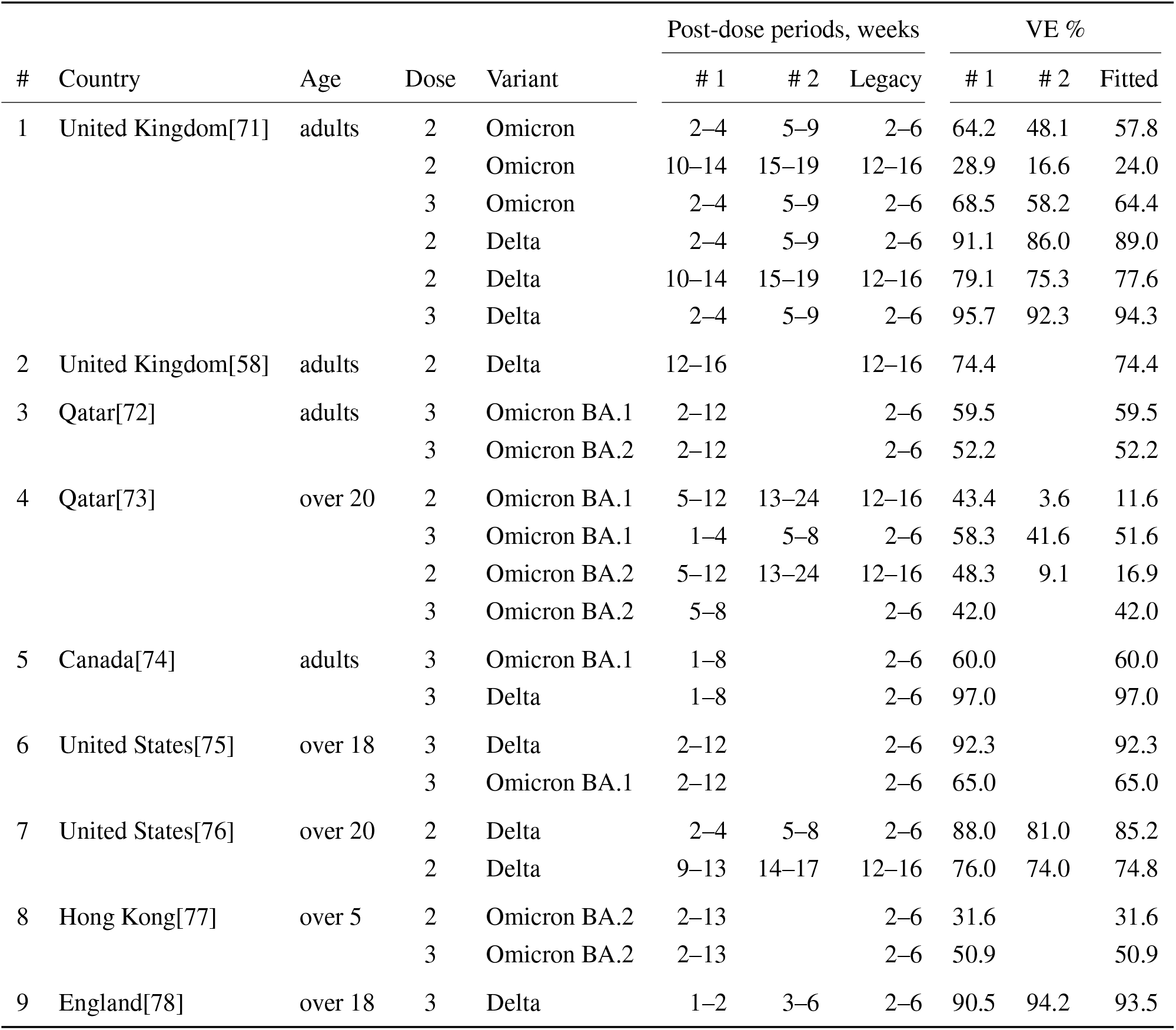
Comirnaty VE estimates against Delta or Omicron. The estimates are shown for study post-dose periods closest to a Legacy post-dose period. If the latter overlapped with two study periods, we combined the two VE estimates into a single VE used in fitting.

**Figure 5:**
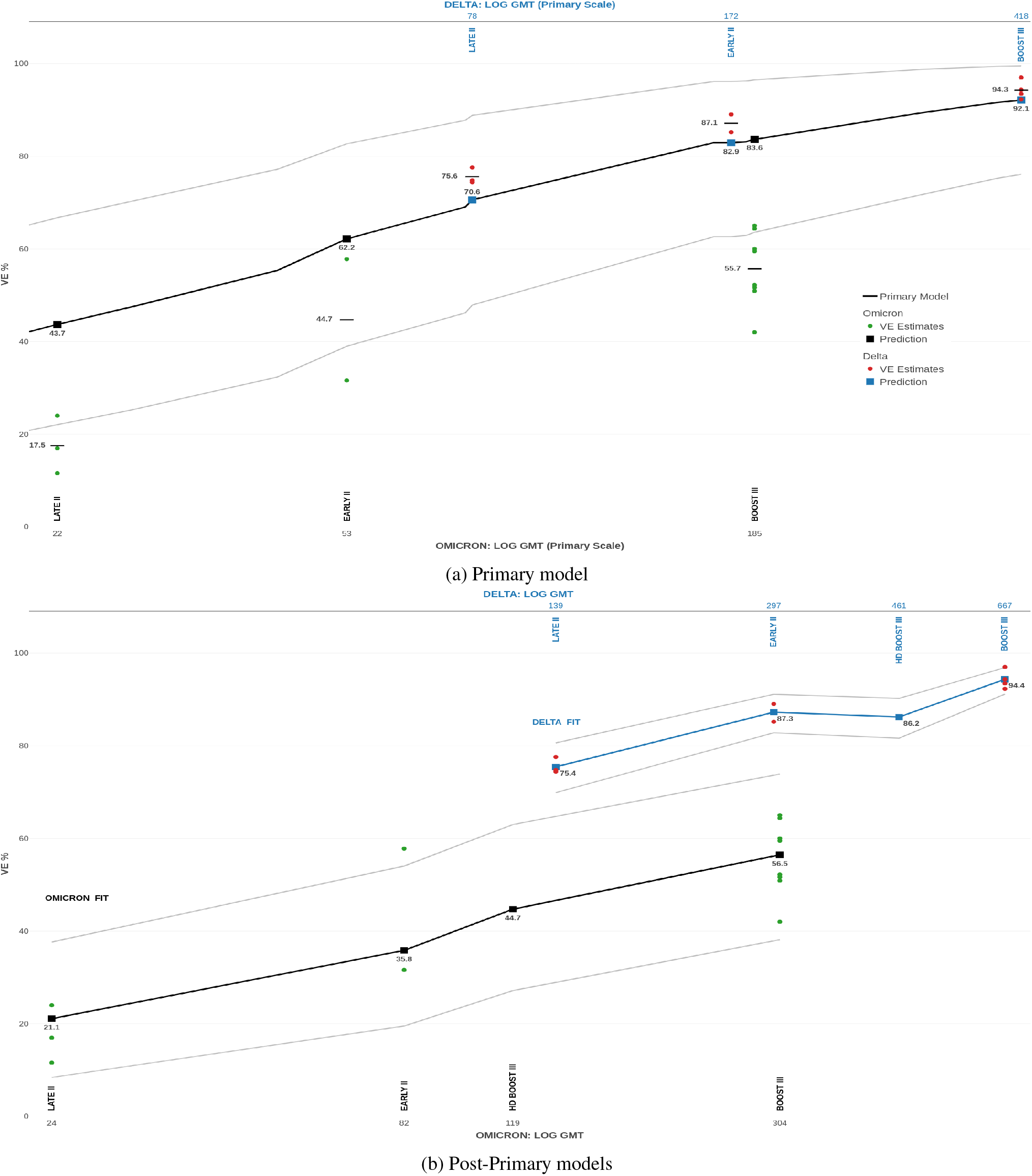
Comirnaty VE for Early group 2–6 weeks after dose II; Late group 12–16 weeks after dose II; Booster group 2–6 weeks after dose III. The top and bottom GMT axes in a plot have the same scale. (a) The Comirnaty model is fitted to “Primary” Legacy NAbTs in seronaive participants. The predictions are based on “Post-Primary” NAbTs[17] divided by 1.51 for Delta and 1.6 for Omicron. (b) Separate models for Comirnaty VE against Delta and Omicron fitted to NAbTs from Post-Primary Legacy. Also shown is predicted VE in haemodialysis (HD) patients 4 weeks after a Comirnaty booster.

We next use only the “Post-Primary” NAbTs and the VE estimates in Table 6 to fit a Post-Primary Comirnaty VE model (2) separately for Delta and for Omicron. Figure 5b shows a good resulting fit for either model. (The predicted VE for Delta in the Early group exceeds the two-dose VE of 82.3% from the Primary model, which is reasonable given the different post-dose periods.) The predicted VE four weeks after boosting haemodialysis patients[19] is 86.2% (95% CI: 81.6–90.2) for Delta, and 44.7% (95% CI: 27.1–63.0) for Omicron, which is respectively 8.8% and 11.8% less than for recently boosted healthy adults. As expected, NAbTs in a given group are substantially lower for Omicron. However, similar NAbTs correspond to a much lower VE against Omicron than against Delta, with the model-specific 95% prediction bands not overlapping. For example, Booster NAbTs against Omicron are similar to the Early NAbTs against Delta — with the GMT of 304 (95% CI: 240–364) versus 297 (95% CI: 265–332) — whereas the VE prediction is 56.5% (95% CI: 38.1–73.9) versus 87.3% (95% CI: 82.8–91.1). To explore this further, we evaluate both models at the Omicron NAbTs per group and simulate prediction differences from the corresponding beta distributions. Again, there are marked differences between the models; see Table 1.

## Discussion

We modelled Comirnaty VE and Vaxzevria VE against the D614G, Alpha, Beta, Delta and Omicron BA.1 variants, and predicted VE for various vaccine doses, immunogenicity cohorts, data subsets and targeted populations. The predictions were substantiated by empirical VE estimates, which further supports NAbTs as correlates of Covid vaccine protection. The joint modelling of both vaccines suggests that the same CoPs could apply across the mRNA and adenovirus-vector vaccine platforms. The deviations of the “leave-one-variant-out” predictions of VE against Alpha, Beta or Delta were between 2% and 25% lower than the corresponding deviations in the state-of-the-art paper[10], while the 95% prediction bands obtained here were more realistic. The joint modelling of the pre-Omicron variants suggests their similarity in VE aspects, such as neutralisation mechanisms and disease symptoms. A pre-Omicron model over-predicted VE against Omicron by between 18% and 28%, and a separately-fitted model was necessary. Depending on the participants group, identical NAbTs corresponded to between 18% (95% CI: 1–33) and 31% (95% CI: 13–50) lower predicted VE against Omicron than against Delta; hence, a GMT fold for these variants would underestimate the drop in protection against Omicron. A separate VE model and CoPs could therefore be needed for each set of variants/subvariants — such as the four pre-Omicron variants or, potentially, Omicron and its subvariants — that are similar in VE aspects. (Alternative biomarkers, such as 80% NAbTs, and/or models, such as fixed-effect models, might suit joint VE modelling with dissimilar variants.) We also note a markedly reduced Comirnaty VE of 45% (95% CI: 27–63) against Omicron BA.1 in recently boosted haemodialysis patients, which might also happen in immunocompromised and other disease subgroups.

The data-driven frameworks[9, 8, 10, 11] combine NAbTs from studies with different vaccines, participants and protocols by normalising vaccine-induced NAbTs by the geometric mean of unvaccinated convalescents’ NAbTs. This makes data from several studies available, and a biased study might be compensated by other studies. However, convalescent NAbTs are added per immunogenicity study, thus increasing the model error. Age-dependent biases can emerge, as younger convalescents have lighter Covid and lower NAbTs, whereas younger vaccinees have stronger immunity and higher NAbTs, than do older convalescents[27] and vaccinees, respectively. Between-study errors can remain high after normalisation. Study populations can be small and unrepresentative: for example, the mRNA-1273 study[28] only had three convalescents and 14 vaccinees. The modelling[9] combined Wild-type NAbTs with efficacies estimated for D614G; as the latter NAbTs can be lower — for instance, by 2.3 folds (95% CI: 1.9–2.6) [15] — this could increase the model biases. To model VE against a variant its GMT fold versus the ancestral Wild Type is estimated for each vaccine in an immunogenicity study, with these estimates averaged across the vaccines and studies[10, 11]. A “variant” VE model is defined by rescaling the ancestral model[9] by this average fold. To predict Comirnaty VE, the variant model is evaluated at the GMT fold of Comirnaty NAbTs versus convalescent NAbTs, both against the Wild Type. Consequently, the variant folds increase uncertainty; ancestral model errors transfer to each variant model; and variant-specific data are underused. (The predictions, in Figure 3b, of two-dose Comirnaty VE from D614G data suggest that ancestral variants are informative in some cases.) The initial immunogenicity studies, underpinning the ancestral model[9], provided only limited data on age subgroups: for example, a Comirnaty study[29] had 11 vaccinated participants aged 18–55 and 12 aged 65–85. Predictions[11] rely on these small samples and do not cover age bands besides the original. These studies provided little-to-no data on disease subgroups, whereas later studies may omit ancestral variants and/or convalescents altogether[18, 19]. Furthermore, if convalescent NAbTs come from the same subgroup as vaccine-induced NAbTs then subgroup factors, such as kidney disease and older age, can distort the subgroup’s normalised NAbTs relative other normalised NAbTs used, which come from a general population.

The proposed VE framework uses vaccine-induced NAbTs from a large study, similarly to the approach[30], and/or from coordinated studies with compatible outputs. Errors from convalescent normalisation and from laboratory differences are reduced or eliminated, whereas a large study population can represent wider populations adequately and also facilitate covariate matching. (Although inputting Post-Primary Legacy NAbTs into a Primary model increased the uncertainty, this effect is localised, compared to that of convalescent normalisation of every model. Furthermore, coordinated studies could be designed to be compatible eg, by including between-study controls.) Similar variants/subvariants are modelled jointly, thus better utilising information, and any number of them can be so modelled. Uncertainty due to variant folds is absent, and models are easily updated with new data. Predicting VE in subgroups is straightforward, and reasonably-large samples can be available, as with age subgroups of Legacy. Unlike other CoP frameworks, we incorporate one-dose VE, which covers once-vaccinated subgroups and improves fitted models, especially at low and mid-range NAbTs. We also add new techniques, such as modelling VE with a beta distribution; imputing censored NAbTs with a simple algorithm; bridging NAbTs from different studies by using a fitted VE model; and weighting VE components by SARS-CoV-2 genotype incidence.

The proposed framework has its limitations. For example, incompatible NAbT datasets are excluded from modelling. Predictions are unavailable for vaccines, variants or subgroups absent from compatible immunogenicity studies. Even a large immunogenicity study can have biases, and its protocol and participants may change between study occasions. Models for a divergent variant may require VE estimates against it from observational studies. Then we may not speed up VE assessment for general populations, but could for subgroups, subsequent boosters and subvariants. Crucially, data-driven VE frameworks, including[8, 9, 10, 11], rely on statistical associations between NAbTs and VE estimates, with both obtained in distinct populations and mainly without randomisation. These populations could differ due to health and demographic factors; vaccination characteristics, such as post-dose times; prevalence of SARS-CoV-2 genotypes; differently-defined symptomatic VE between studies[31]; and the variation in Covid severity and symptoms across variants. Fitted VE models can therefore be biased and/or extrapolate poorly to novel situations. As data-driven VE predictions sidestep causality, they can be difficult to verify: even definitive CoPs from a randomised controlled trial are incomplete — NAbTs mediated 69% of mRNA-1273 efficacy against the Wild Type[3] — and especially so for vaccines and variants not in the trial. These challenges could be met by advancing VE modelling methodology, and by updating and validating fitted models with historic and recent data. Reassuringly, our framework meets several heuristic conditions[32] for a causal dependence of VE on NAbTs. First, the framework is empirically substantiated in different populations and modelling problems. Second, the fitted models exhibit a strong dose-response relationship, with higher NAbTs mostly corresponding to higher VE; see Figure 1. Third, mediation by NAbTs of Covid vaccine protection is biologically plausible and is supported by experimental and modelling evidence[2, 3, 4, 8, 9]. Despite the differences between the Legacy and VE study populations the former population captured the common biological processes and appropriately modelled immune responses in the latter. In general, however, population differences could matter, especially to VE prediction in subgroups.

Extending our framework with covariate matching could compensate for population differences, as well as facilitate individual predictions for a country or VE study population. We could accommodate additional VE types[9, 10, 11], VE modelling over continuous time and with alternative biomarkers, such as 80% NAbTs and metrics based on T-cells or on binding antibodies[8]. Statistical enhancements could include meta-regression, extra covariates, censored data algorithms and provisions for uncertainty in NAbTs, and benefit from Bayesian techniques, such as prior distributions and advanced posterior simulations. Enhancements could also draw on alternative VE frameworks: for example, genetic distances[7] could help identify variants for separate modelling. Data-driven VE frameworks could be compared under various assumptions, prediction tasks, practical conditions and available datasets. Alternative frameworks could be combined to support ensemble predictions and scenario analyses. Data-driven frameworks could be validated and improved with data and methods from definitive CoP frameworks[33, 2, 5, 6, 4].

As immunogenicity and VE data are crucial, enhancing them could improve VE predictions. In particular, the definition of VE could be standardised across observational studies and finer detail provided on study populations, especially on subgroups. Our results and those in [10, 9, 30] demonstrate the benefits of using comparable immunogenicity data, such as those obtained by the Legacy[15, 16, 17] and the haemodialysis studies[18, 19]. Uniform protocols, standardised external and internal controls could make NAbTs more comparable across immunogenicity studies and between study occasions. Study designs, including enrolment criteria, could deliberately support VE prediction and pandemic decisions more generally. Immunogenicity studies could be coordinated to cover key subgroups, vaccines and vaccine regimens, and to run comparable, rapid and informative assays with emerging variants/subvariants. Such studies could save resources; enable accurate, real-time VE predictions; and support fast, precisely-targeted measures against pandemic threats.

Ultimately, the studies and modelling frameworks that we discussed are tools for reducing human and economic costs of Covid-19. The proposed framework can be a useful additional tool, especially at this stage with the fast-evolving virus and limited appetite for pandemic measures and spending. It could also provide a VE prediction blueprint for tackling future epidemics.

## Data Availability

All data used for the paper are publically available online.

https://github.com/XitificOS/prediction_Covid_VE

## Competing interests

The authors declare no competing interests.

## Funding

This work received no external funding.

## Data availability

Data and R code used for this paper are freely available from the paper's GitHub repository

## Acknowledgments

O.V. is grateful to R. Bailey, I. Bartlett, J. Briggs, J. Fogaros and A. Langley for their support and encouragement during this research. The authors thank R. Lodwick, B. Bogacka and S. Bird for their suggestions which substantially improved the paper, and to A. Kajava for fruitful discussions. We are grateful to the authors of Gilbert et al.[33], Khoury et al.[9], Cromer et al.[10], Wall et al. [15, 16] and Carr et al.[18] for making their data and programming code publicly available, which greatly facilitated our research. We thank D. L. Bauer for providing additional data from the Crick Legacy Study.

## A Methods

### A.1 Immunogenicity studies with SARS-CoV-2 variants

Immunogenicity data used in our modelling were obtained in the ongoing Legacy SARS-CoV-2 Immunogenicity Study, run in the UK by the University College London Hospitals (UCLH) and the Francis Crick Institute[15, 16]. The study participants — Comirnaty or Vaxzevria vaccinees — are healthcare staff from UCLH and research staff from the Crick Institute.

#### “Primary” Legacy Study

This study investigated the primary vaccination with Comirnaty or Vaxzevria. The Comirnaty stream included a one-dose cohort with 186 participants and a two-dose cohort with 160 participants, with 248 unique participants overall, some of whom in both cohorts. The corresponding numbers in the Vaxzevria stream were 50, 63 and 106. Study demographics and other details can be found in the references[15, 16], with anonymised data available at [the GitHub repository for](https://github.com/davidlvb/Crick-UCLH-Legacy-AZ-VOCs-2021-06) [16]. Gender, Ethnicity and BMI were not significantly associated with the NAbTs, while the factors that were are as follows. Participants’ Age, reported in 5-year bands, is between 20 and 70, with a median of 42, for Comirnaty, and between 20 and 65, with a median of 32, for Vaxzevria. For either vaccine, about 28% participants had Prior Covid sometime before study visits. Time since vaccination in the one-dose cohort was 2–9 weeks, with a median of 4, for Comirnaty and 2–12 weeks, with a median of 6, for Vaxzevria. Time since the second Comirnaty dose was 1-9 weeks, with a median of 4 and 1–26 weeks, with a median of 4, for Vaxzevria. Between-Dose Interval was 2–11 weeks for Comirnaty and 6–14 weeks for Vaxzevria, with a median of 9 for either. The study investigated an ancestral SARS-CoV-2 strain with the D614G mutation and the Alpha, Beta and Delta variants. The Comirnaty and Vaxzevria investigations occurred three weeks apart under identical experimental protocols and output comparable data[15, 16].

#### “Post-Primary” Legacy Study

A follow-up study[17] six months later investigated immunogenicity after two or three (ie booster) doses against the Alpha, Delta and Omicron BA.1 variants. The Comirnaty stream included 199 participants 2–6 weeks after dose two; 136 participants 12–16 weeks after dose two; and 85 participants 2–6 weeks after a booster. Some of the participants were shared between these groups[17] and also with the “Primary” Study. The demographics of the twice-vaccinated groups were similar in both Legacy studies. The boosted participants were older, with the median age/IQR of 53[45–59].

#### Studies in patients on haemodialysis

Two immunogenicity studies were done by the UCLH and Crick in Comirnaty-vaccinated patients on haemodialysis. The first study[18] investigated the same doses and variants under the same protocols as in “Primary” Legacy. We use data from 55 twice-vaccinated, initially seronaive patients, whose mean (standard deviation) age was 60(12), and 37% of whom were females. The second study[19] investigated immunity against Delta and Omicron BA.1 after second and booster doses, under the same protocol as in “Post-Primary” Legacy. We used data from 84 participants, who were demographically similar to those in the first study; their prior seropositivity status was not published.

#### Neutralising antibody titres

Each participant’s plasma was individually tested per dose and variant at the 40, 160, 540 and 2560 titres set in duplicates. Automated in-vitro neutralisation assays measured the proportion of a live-virus plaque neutralised at a pre-set antibody titre. A titre corresponding to 50% neutralisation was estimated per test by fitting a four-parameter logistic model. We abbreviate these estimates as NAbTs and omit 50% for simplicity. Any NAbT below 40 was less than the assay limit of detection (LOD) and recorded as 10; any above 2560 was greater than the LOD and recorded as 5120[15]. When no neutralisation was observed, a NAbT of 5 was recorded. The resulting distributions of log_10_-transformed NAbTs are illustrated for Comirnaty in Figure 6. The Normal-distribution assumption is noticeably violated, especially after the first dose.

**Figure 6:**
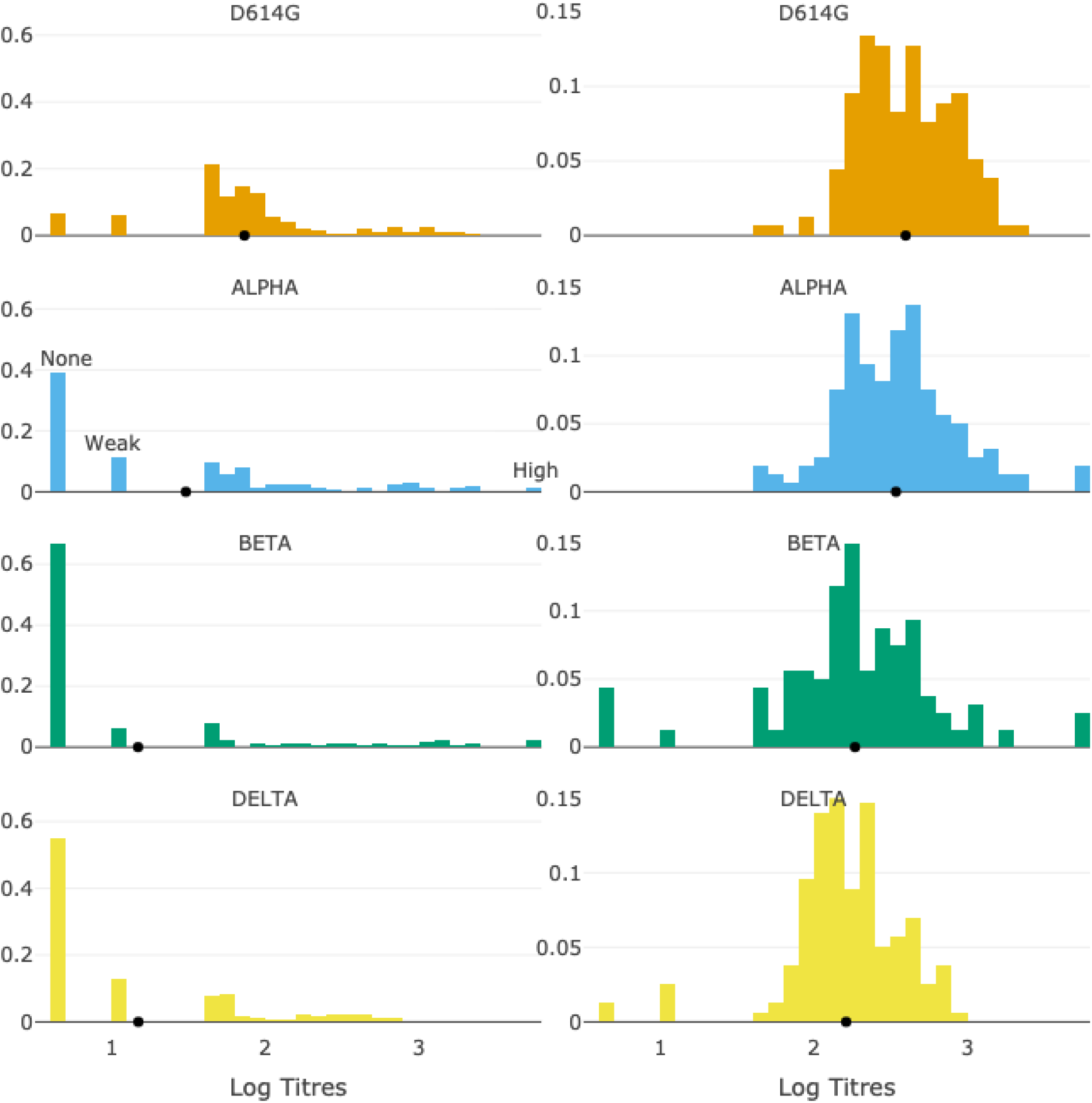
Distributions of log_10_-transformed NAbTs after one (Left) and two (Right) Comirnaty doses. Each distribution’s geometric mean titre (GMT) is (•). Assay LODs are illustrated for Alpha after one dose: “None”, recorded as 5, means no observed neutralisation; “Weak”, recorded as 10, means an estimated NAbT below 40; “High”, recorded as 5120, means an estimated NAbT above 2560. Data source[15].

### A.2 Modelling

#### Imputation of censored titres

Many of the Legacy NAbTs were censored, especially left-censored. For example, among Comirnaty NAbTs against Beta 72% were below and 2% above the LOD after one dose versus 6% and 3% after two doses; for Vaxzevria, these percentages were 68% and 4% after one dose versus 40% and zero after two doses. To mitigate this censoring, we developed an imputation algorithm as follows; see Figure 7. First, we fit a parametric distribution to the log_10_ NAbTs, per dose and variant, with the *fitdistrplus* R package for censored data. Next, we compute representative log_10_ NAbTs for censored titres below log_10_(40) in the distribution’s left tail, and above log_10_(2560) in the right tail. These representative NAbTs are defined per tail as

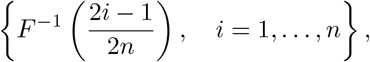

based on [34, p. 19]. Here, *F*^*−*1^(*·*) is the inverse c.d.f. of the fitted distribution, and *n* is the number of censored titres in the tail. This imputation can underperform with too few uncensored NAbTs, and is optional but should be used consistently in joint vaccine modelling, defined below.

**Figure 7:**
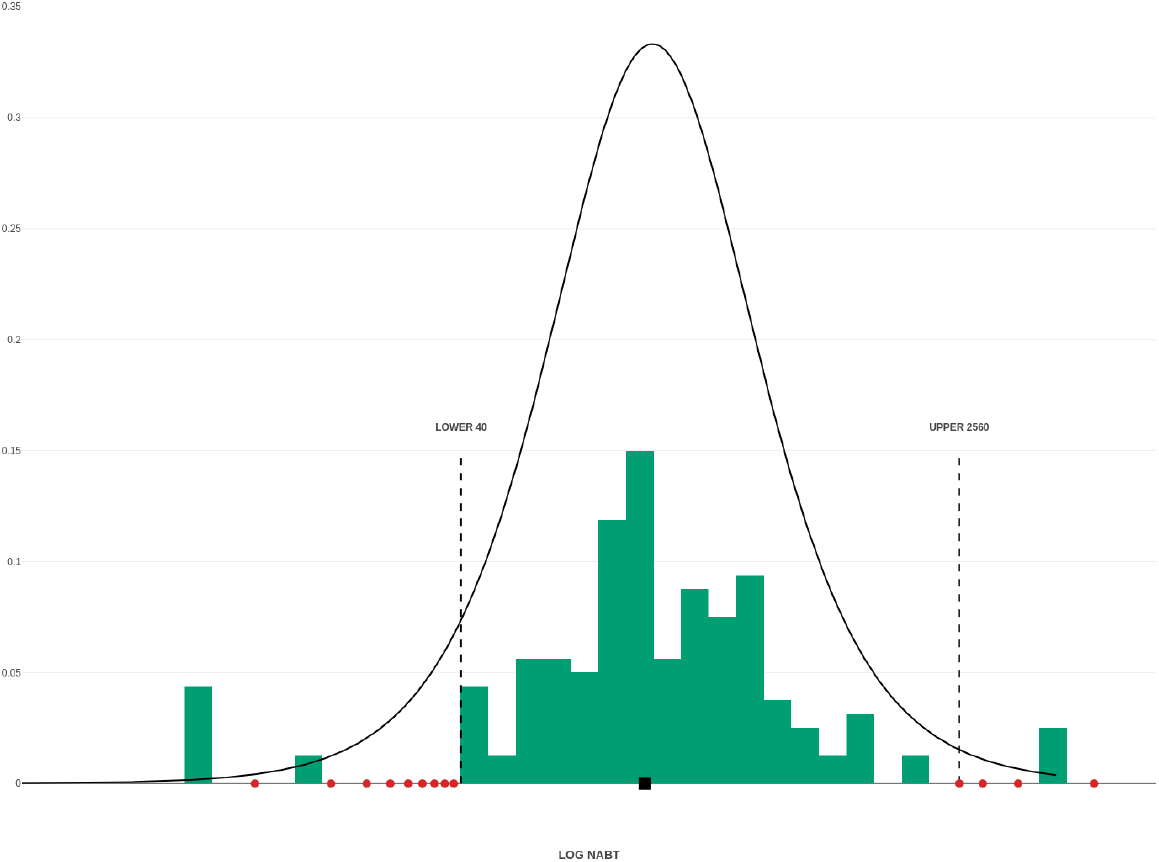
Imputation of NAbTs against Beta for two Comirnaty doses. Logistic pdf was fitted to the censored NAbTs. Nine values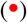 were imputed below the LOD, and four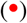 above the LOD. GMT is shown(■)

#### Symptomatic effectiveness of a vaccine

We model the symptomatic effectiveness (VE) of a vaccine dose against a variant/subvariant for the given population and post-dose period. Applied to Vaxzevria and Comirnaty in this paper, this modelling could generalise to other Covid vaccines and VE types. To model VE we fit a function (defined shortly) of NAbTs to symptomatic VE (or efficacy) estimates from vaccine observational studies and clinical trials; see Extended Data. The assumed post-vaccination period for VE of one dose is from 7 days post-dose until dose two, and for VE of two doses, it is from 7–14 days post-dose until 3–5 months later; these are roughly the periods behind the VE estimates used; other periods are assumed as indicated. *Direct* modelling for a vaccine uses VE estimates and NAbTs for the vaccine; *indirect* modelling uses a model fitted with data on one vaccine to model the VE of another vaccine, by evaluating this model at the NAbTs of the latter; *combined* modelling fits a model function to VE and NAbTs for both vaccines. The two joint modelling approaches are useful when many fewer VE estimates are available on one vaccine than on another. To validate our modelling, we obtain “leave-one-variant-out” predictions: a model, fitted without data on a variant, predicts VE against the variant by evaluating the model at the variant’s NAbTs. These predictions apply, for example, to situations when NAbTs, but not VE estimates, are available for a new variant/subvariant; note an analogy to the “leave-one-vaccine-out” validation[8, 9].

Following refs[35, 36] we first define the *individual* protection function for the probability that a person vaccinated with given doses is protected over the assumed period against symptomatic Covid caused by a given variant/subvariant:

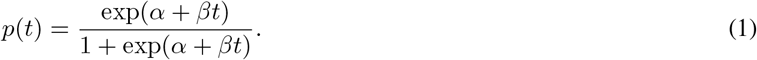

Here, the covariate *t* is a log_10_-transformed 50% neutralising antibody titre (NAbT) against the variant, and *α* and *β* are the model parameters to estimate. The percent of protected vaccinees, from a population of size *N*, is modelled by the *population* protection function

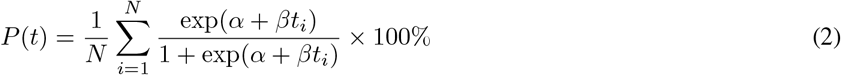

Here, *t*_*i*_ is the log_10_ NAbT in person *i*, with all *N* titres corresponding to a particular vaccine, dose, variant/subvariant and time period. This function is an expectation, with respect to distributions of log_10_ NAbTs, of individual protection functions. We apply this population function, also used in[35, 37, 9, 10], as the model for VE. If censored NAbTs are imputed, this function is evaluated at original non-censored titres and at the imputed “representative” titres; otherwise, at the originally-reported NAbTs, including 5, 10 and 5120. The function (2) is mathematically equivalent to a standard definition of VE, if the VE study population is seronaive: that is, has no antibodies due to SARS-CoV-2 infection. This equivalence, easily seen from the VE definition in [35], is approximate if infection-induced antibody levels in the population are low. If these levels were high, the function can still model VE adequately but is no longer equivalent to the formal VE definition. Many VE studies exclude identifiable prior Covid-19 cases or adjust for them, so as not to bias VE estimates[38, p. 27].

#### Approximating the VE model to aid visualisation

Since the model function (2) inputs multiple NAbT samples, each of which multidimensional (see Figure 6), its plotting and visualisation are challenging. We therefore plot the function versus log_10_ of the geometric mean titre (GMT) per NAbT sample. To determine whether the resulting plots are realistic, we approximate this function as follows. Each individual NAbT in a sample is assumed to equal the sample’s GMT, in which case (2) is equivalent to the individual function (1). Hence, we define an approximate VE model based on a function of GMT only, as in [8]. Figure 8 illustrates this approximation for Comirnaty: the greatest difference is about −2% for one-dose VE against Alpha, and both sets of the 95% prediction bands (only shown for the population model) are close. This close approximation suggests some modelling robustness to the shape of NAbT samples, provided their GMTs are stable.

**Figure 8:**
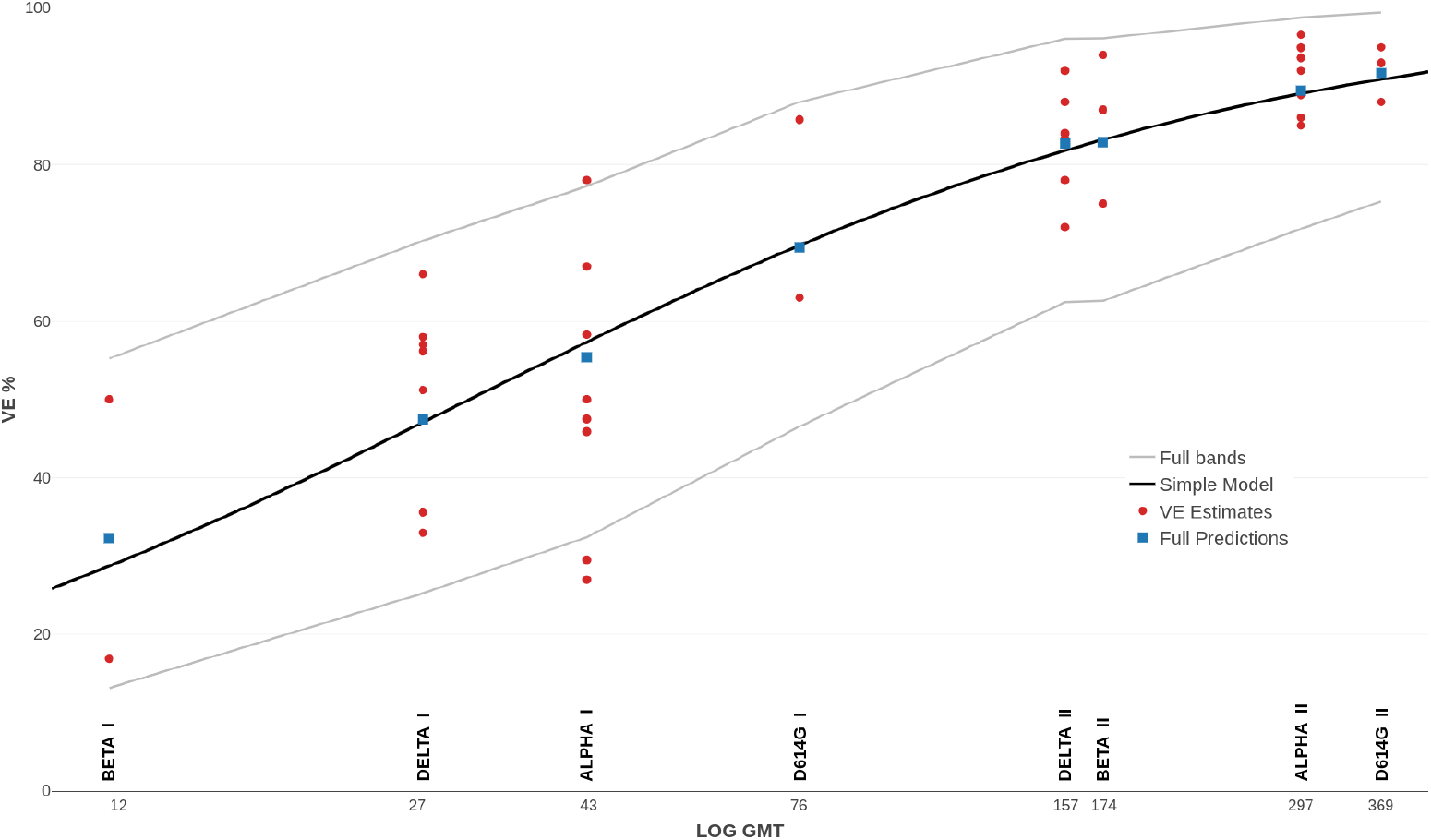
Simplified versus standard models for Comirnaty. The fitted standard model is the same as in Figure 1a. The 95% prediction bands (not shown) for the simplified model are close to those for the standard model.

**Figure 9:**
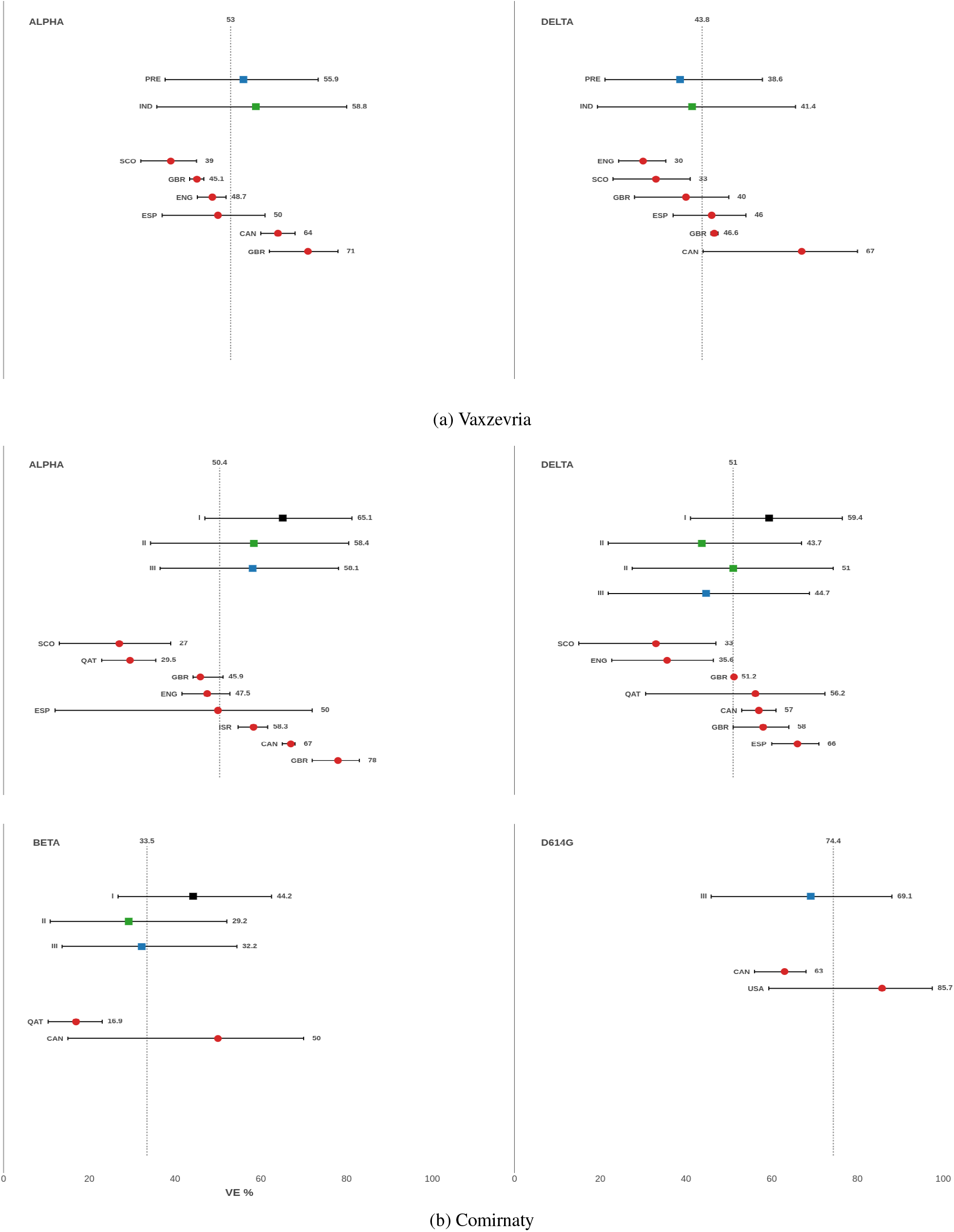
Predictions of one-dose VE against a variant versus VE estimates from indicated countries; see Tables 2 and 3. (a) “Leave-one-variant-out” predictions of Vaxzevria VE, including PRE direct predictions from Vaxzevria data; IND indirect prediction from a Comirnaty-fitted model at Vaxzevria NAbTs. (b) Predicted Comirnaty VE. Epoch I: predictions from D614G data. Epoch II: predictions from D614G and another variant; Epoch III: predictions from three other variants, ie, “leave-one-variant-out” predictions.

**Figure 10:**
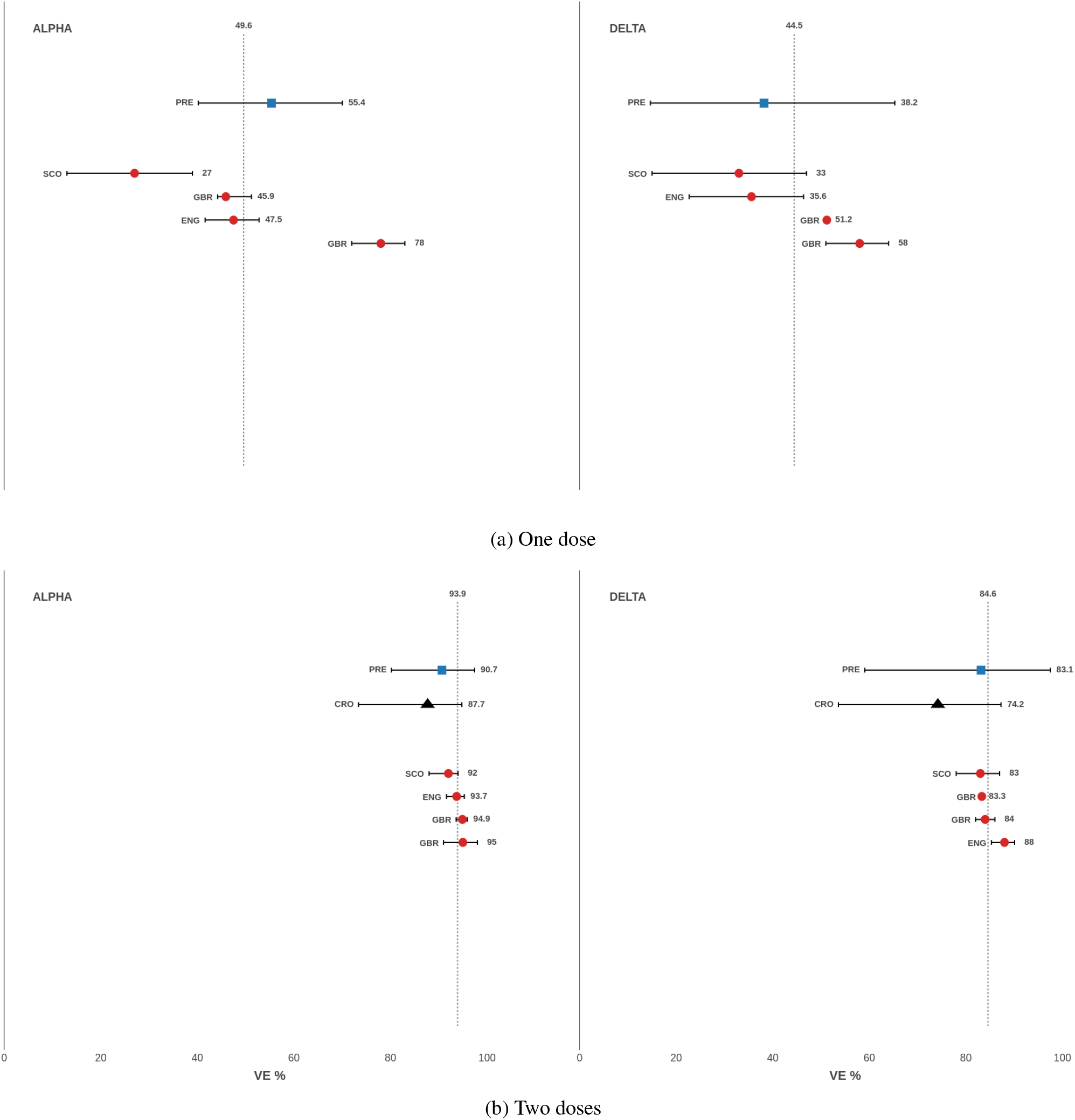
“Leave-one-variant-out” predictions of Comirnaty VE against Alpha or Delta in the UK, from UK data only. PRE obtained predictions; CRO predictions[10], which are not country-specific.

#### Combined VE against multiple variants

The assumed VE studies in diabetes and kidney disease subgroups estimated VE for the prevalent SARS-CoV-2 variants combined; see Table 5. We therefore predict a combined VE for each of these studies individually. This VE is defined as *P* (*t*) = *s*_*k*_*P*_*k*_(*t*), where *P*_*k*_(*t*) is the model (2) fitted for variant *k*, and *s*_*k*_ is the variant’s share of the Covid cases in the study country within the study dates. To compute *s*_*k*_, we use data from *covariants*.*org*[39] on country-wise weekly counts of genetic sequences reported to the [GISAID database](https://gisaid.org/) for specific variants. These counts are combined with Covid cases, reported by [Johns Hopkins University CSSE](https://github.com/CSSEGISandData/COVID-19/tree/master/csse_covid_19_data/csse_covid_19_time_series) per week and country to approximate weekly case counts per variant, so as to account for varying sequencing rates over time. The share *s*_*k*_ is based on these weekly case counts summed over study duration. As the assumed VE studies started when the D614G mutation was prevalent worldwide, we lump any ancestral sequences as the D614G “variant”. The other dominant variants were Alpha, Beta or Delta; we ignore the very few cases of Gamma, as this variant was not examined by Legacy.

#### Modelling when NAbTs are not directly comparable between studies

Suppose we predict Comirnaty VE against Delta from a Comirnaty model fitted to “Primary” Legacy NAbTs by evaluating it at “Post-Primary” Legacy NAbTs against the variant. As some participants and experimental protocols differed, the two studies produced somewhat incomparable NAbTs. To resolve this we first refitted the “Primary” Comirnaty model by excluding participants with post-second-dose time outside 2–6 weeks, which was the corresponding period in the “Post-Primary” study. (We only included NAbTs from seronaive participants of both studies.) The Early “Post-Primary” group with 148 participants 2–6 weeks after dose two closely matches the corresponding “Primary” group with 96 participants: the median age is 42 [IQR: 32, 52] in both and the female share is 66% and 64%, respectively. Given the groups similarity, if Delta NAbTs were measured in this “Post-Primary” group instead of the original “Primary” cohort, the resulting fitted model would have been close to the original “Primary” model. To compare Post-Primary NAbTs, denoted ***t***^*Post*^, against Delta with Primary NAbTs, denoted ***t***^*Prime*^, we find *κ* such that *P(****t***^*P ost*^*/κ)* = *P(****t***^*Prime*^), where *P* (*·*) is the original “Primary” model (2). To predict VE against Delta in the Post-Primary groups, we divide their NAbTs by this *κ*. We do not account for uncertainty in *κ*, which could be done by simulations, such as bootstrap. A similar *κ* can be found for each variant investigated in both studies. In particular, *κ* for Delta, 1.51, was close to that for Alpha, 1.75, which indicates that *κ* is mainly due to protocol and population differences. To predict VE against a variant, such as Omicron, that is absent from the “Primary” study, we find *κ* as a compromise between the two variants. Specifically, we used *κ* = 1.60 that minimises the maximum prediction error of VE against Alpha or Delta, when the corresponding Early Post-Primary NAbTs are inputted into the Primary model.

#### Statistical modelling

We model covariate-adjusted estimates of VE directly, by fitting the model function (2) and assuming that VE follows a beta distribution, which is suited to interval-bounded outcomes. We assume VE strictly between 0% and 100%, and exclude the two bounds for simplicity; they can be handled with an extended beta distribution. Since VE estimates come from different studies and have unequal variance, a meta-regression would be necessary in theory. To our knowledge, however, no open-source software existed for nonlinear, or even linear, meta-regression with beta distributions. Instead of developing custom software, we adopt standard beta regression, as follows. The function (2) is fitted by nonlinear beta regression under the parametrisation proposed in [40, 41], which fits the model parameters *α* and *β* together with a precision parameter *φ*. The corresponding log-likelihood function is maximised numerically with the *optim* procedure in R. To obtain a starting guess we fit the individual function (1) by using log_10_-transformed geometric mean titres (GMT) per NAbT sample. Since a logit transformation of (1) is *α* + *βt*, this fitting applies a generalised linear model framework extended to beta regression, within the *betareg* package [41]. The 95% prediction interval is defined with 2.5% and 97.5% quantiles of the beta distribution output by regression. (To our knowledge, beta regression has not been used before to model vaccine efficacy or effectiveness.)

We explored the following analysis options to NAbTs data: (1) Include or exclude prior Covid cases from the Legacy data; and (2) Impute censored NAbTs or not. Excluding prior Covid cases is theoretically attractive, since the model (2) is then equivalent to the VE definition. A downside is the discarding of data, for example, up to 28% NAbTs from Primary Legacy[15]. With many NAbTs, as available for Comirnaty, the resulting predictions were similar across options — for example, the two-dose “leave-one-variant-out” predictions in adults differed by 1% at most. With relatively few NAbTs, as for Vaxzevria, some predictions differed substantially. To avoid cherry-picking of data, we set the same options in related prediction tasks, such as the Primary modelling of Comirnaty. When required, we select subsets of NAbTs to match populations, for example, by Age and Time post-dose. Tables 7 and 8 list the settings applied in this paper. Alternative settings can be explored with the provided software package.

**Table 7:** Primary models for VE against pre-Omicron variants. All NAbTs from Primary Legacy[15, 16] and the VE estimates from Tables 2 and 3 were used. The NAbT sample size, after I or II doses, is averaged per variant; data on four variants was used for Comirnaty and three (without Beta) for Vaxzevria. Participants with prior Covid were included (Y) or excluded (N); censored NAbTs were imputed (Y) or not (N). †Included twice-vaccinated participants had 2–6 weeks after dose two, to match the twice-vaccinated participants of Post-Primary Legacy[17]. The models are shown in Figure 1 and† Figure 5a.

| Model | Parameter Estimates (SE) |  |  | Sample Sizes: NAbTs after and VE |  |  | Data Options |  |
| --- | --- | --- | --- | --- | --- | --- | --- | --- |
| | $\alpha$ | $\beta$ | $\phi$ | Comirnaty I/II | Vaxzevria I/II | VE | Covid | Imputed |
| Comirnaty | -3.5 (3.1) | 2.4 (1.7) | 17.4 (24.4) | 132/111 |  | 43 | N | Y |
| Vaxzevria Indirect | -2.3 (2.3) | 1.8 (1.3) | 16.5 (23.1) | 184/159 |  | 43 | Y | N |
| Vaxzevria Direct | -3.2 (3.5) | 2.5 (2.6) | 24.5 (34.0) |  | 49/63 | 25 | Y | N |
| Combined | -3.3 (3.4) | 2.3 (1.9) | 18.5 (25.8) | 184/159 | 49/63 | 68 | Y | Y |
| Comirnaty† | -3.4 (3.0) | 2.3 (1.6) | 17.4 (24.4) | 132/97 |  | 43 | N | Y |

**Table 8:**
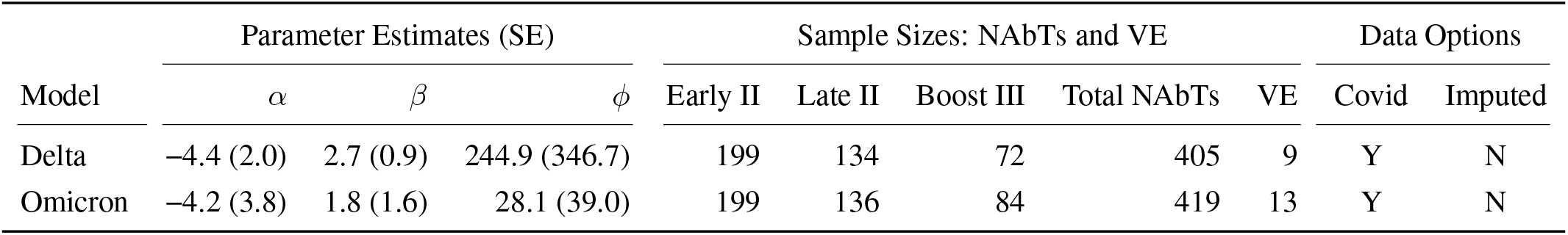
Primary Comirnaty models were separately fitted for Delta or Omicron by using the corresponding two or three-dose NAbTs from Post-Primary Legacy[17]. VE estimates, from Table 6, match the post-dose periods. See Figure 5b.

| Model | Parameter Estimates (SE) |  |  | Sample Sizes: NAbTs and VE |  |  |  |  | Data Options |  |
| --- | --- | --- | --- | --- | --- | --- | --- | --- | --- | --- |
| | $\alpha$ | $\beta$ | $\phi$ | Early II | Late II | Boost III | Total NAbTs | VE | Covid | Imputed |
| Delta | -4.4 (2.0) | 2.7 (0.9) | 244.9 (346.7) | 199 | 134 | 72 | 405 | 9 | Y | N |
| Omicron | -4.2 (3.8) | 1.8 (1.6) | 28.1 (39.0) | 199 | 136 | 84 | 419 | 13 | Y | N |

## B Extended data: estimates of symptomatic VE

### Pre-Omicron variants in adult populations: primary vaccination

We selected published studies of symptomatic effectiveness (or efficacy) of Comirnaty or Vaxzevria against a specified SARS-CoV-2 variant or if the predominant variant was evident from study dates and locations. Primary VE models were fitted to the reported covariate-adjusted estimates of VE in general adult populations, aged 18 and above. The estimates of one-dose VE corresponded to 7–14 days after the dose until dose two, which was administered 4–6 weeks later. The estimates of two-dose VE corresponded to a period 7–14 days after dose two until 3–5 months after. These early-pandemic studies did not stratify VE by post-dose periods. Notably, later studies of VE stratified by post-dose periods, reported stable two-dose VE up to 4–5 months with pre-Omicron variants; see Section 3 of [42].

For Comirnaty, we included 43 VE estimates from 15 studies; see Table 2. Study 11 is the original Pfizer-BioNTech Phase 2/3 vaccine trial at 130 sites in the US and 22 elsewhere. It reported efficacy estimates for a period between May and November 2020[55], when over 90% of the SARS-CoV-2 infections in the US, and similarly world-wide, were caused by an ancestral strain with the D614G mutation[61]. For this study, we inferred the one-dose VE as suggested in [62] but using revised data from the FDA submission[54] and applying the same Bayesian modelling as applied for two-dose VE by trial investigators [54]. Study 4 reported[46] VE estimates by combining 87% Comirnaty and 13% Moderna vaccinees. These estimates were not outliers and were included. The Comirnaty VE estimate against Beta in Study 8 is conditional on Comirnaty VE against Alpha being 95%; see Table 3[51]. For Vaxzevria, we included 25 estimates of its symptomatic VE from eight studies; see Table 3. The two-dose Vaxzevria efficacy against Beta, estimated as 10.4% [CI95: −76.8, 54.8] in a small trial [63] with 717 placebo and 750 vaccinated participants, was excluded as an extreme outlier. Study 3 re-analysed the original Vaxzevria trial [64] to compute separate VE against Alpha and against an ancestral type, which we inferred to have the D614G mutation. Studies 13 and 7 in Tables 2 and 3, respectively, refer to the same UK study of self-reported Covid symptoms in Vaxzevria or Comirnaty vaccinees. The study was initially reported by[57] for VE against Alpha, followed by [58] for VE against Alpha or Delta. We only used the VE estimates against Alpha from the former paper, as the two-dose Vaxzevria VE estimate of 97% [CI95: 93, 98] in the latter is a likely outlier.

### Pre-Omicron variants in age and disease subgroups

To assess Comirnaty VE predictions in age subgroups (Figure 1d), we used age-specific VE estimates, listed in Table 4, from the same UK study[23]. To assess Comirnaty VE predictions in the kidney disease subgroups (Figure 2) we used the studies and Comirnaty VE estimates listed in Table 5. Note that 20% subjects in Study 5 were vaccinated with the Moderna mRNA-1273 vaccine. As these studies reported combined VE for the prevalent variants, we also state approximate shares of the variants in these studies.

### VE estimates, against Delta or Omicron, in post-Primary Comirnaty modelling

Comirnaty VE estimates, in Table 6, against Delta or Omicron were used to assess predictions from a re-scaled primary Comirnaty model (Figure 5a), and to fit a separate model to either variant (Figure 5b.) Published estimates were available for two or three (booster) doses in general adult populations, except for Study 8 with subjects aged 5 and above. Study 9 reported estimates in the 18–49 and 50+ age groups; as the estimates were very similar, they were combined. The earlier Omicron studies did not specify its subvariant, which we presumed to be BA.1. Although the Legacy study[17] investigated this subvariant, we included VE estimates against BA.2, which are similar[20, 21]. The studied reported VE estimates for specific post-dose periods, which we matched to the post-dose periods in Legacy. If a Legacy period overlapped with two consecutive reported periods, we combined the two VE estimates weighted according to the period’s share in the overlap; see Table 6. (Additional weighting by the estimates variance produced very similar results and was not used.)

## C Supplementary results

**Table 9:** Vaxzevria VE predictions per variant with Primary directly-fitted or Comirnaty-fitted models. Variants: 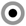 targeted for prediction; 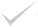 used in model fitting. For each dose cohort, I and II, average NAbT sample size is stated per variant. Prior Covid cases were included; censored NAbTs were not imputed. See Figures 9 and 3 for one-dose and two-dose predictions, respectively.

| Model | Variant |  |  |  | Parameter Estimates (SE) |  |  | Sample Sizes |  |
| --- | --- | --- | --- | --- | --- | --- | --- | --- | --- |
| | D614G | Alpha | Beta | Delta | $\alpha$ | $\beta$ | $\phi$ | NAbT I/II | VE |
| Direct | ✓ | ⊙ |  | ✓ | -3.0 (3.0) | 2.3 (2.3) | 28.2 (39.2) | 48/63 | 14 |
| Direct | ✓ | ✓ |  | ⊙ | -4.0 (5.0) | 3.0 (3.4) | 25.5 (35.5) | 50/63 | 13 |
| Indirect | ✓ | ⊙ | ✓ | ✓ | -2.0 (2.0) | 1.7 (1.2) | 17.5 (24.4) | 183/158 | 24 |
| Indirect | ✓ | ✓ | ✓ | ⊙ | -2.9 (2.9) | 2.1 (1.5) | 15.9 (22.6) | 185/159 | 29 |

**Table 10:** Primary models for Comirnaty VE per variant over three epochs †and for UK only. Variant: 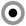 targeted for prediction; 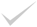 used in model fitting. For each dose cohort, average NAbT sample size is stated per variant. Prior Covid cases were excluded; censored NAbTs were imputed. See Figures 9, 3 and 10 for one-dose, two-dose and UK-only predictions, respectively.

| Epoch | Variant |  |  |  | Parameter Estimates (SE) |  |  | Sample Sizes |  |
| --- | --- | --- | --- | --- | --- | --- | --- | --- | --- |
| | D614G | Alpha | Beta | Delta | $\alpha$ | $\beta$ | $\phi$ | NAbT I/II | VE |
| 1 | ✓ | ⊙ | ⊙ | ⊙ | -2.1 (3.3) | 1.8 (1.6) | 28.1 (39.8) | 130/110 | 5 |
| 2 | ✓ | ⊙ | ✓ | ⊙ | -3.1 (3.1) | 2.2 (1.7) | 15.9 (22.6) | 132/110 | 10 |
| 2 | ✓ | ✓ | ⊙ | ⊙ | -4.0 (3.6) | 2.6 (1.9) | 17.1 (24.2) | 133/111 | 24 |
| 3 | ⊙ | ✓ | ✓ | ✓ | -3.5 (3.2) | 2.4 (1.7) | 16.8 (23.5) | 132/111 | 38 |
| 3 | ✓ | ⊙ | ✓ | ✓ | -3.1 (2.7) | 2.2 (1.5) | 20.1 (28.1) | 131/110 | 24 |
| 3 | ✓ | ✓ | ⊙ | ✓ | -3.5 (3.0) | 2.3 (1.6) | 18.6 (26.1) | 132/111 | 38 |
| 3 | ✓ | ✓ | ✓ | ⊙ | -3.8 (3.7) | 2.5 (1.9) | 15.7 (22.2) | 133/111 | 29 |
| † |  | ⊙ |  | ✓ | -4.1 (2.2) | 2.7 (1.3) | 41.3 (57.9) | 130/110 | 8 |
| † |  | ✓ |  | ⊙ | -4.7 (4.4) | 3.0 (2.4) | 12.6 (17.8) | 134/111 | 8 |

## Notes

### Competing Interest Statement

The authors have declared no competing interest.

### Funding Statement

This study did not receive any funding

### Author Declarations

The study used ONLY openly available human data that were originally located at: https://github.com/davidlvb/Crick-UCLH-Legacy-AZ-VOCs-2021-06 https://github.com/davidlvb/Crick-UCLH-Legacy-Omicron-2021-12 https://github.com/EdjCarr/Crick-HD-AZD-BNT-VOCs-2021-07 https://github.com/EdjCarr/Crick-HD-Omicron-2021-12

